# Bile Acid Metabolism in Multiple Sclerosis is Perturbed and Associated with the Risk of Confirmed Disability Worsening

**DOI:** 10.1101/2025.03.10.25323690

**Authors:** Ida Erngren, Aina Vaivade, Henrik Carlsson, Asma Al-Grety, Torbjörn Åkerfeldt, Ingrid Kockum, Anna Karin Hedström, Lars Alfredsson, Tomas Olsson, Joachim Burman, Kim Kultima

## Abstract

**Background:** Bile acids (BAs) have emerged as important mediators in neuroinflammation and neurodegeneration, important features of multiple sclerosis (MS). This study aimed to examine serum BA levels in newly diagnosed people with MS (pwMS) and explore their association with disability worsening.

**Methods:** The study included 907 pwMS and 907 matched controls from the Swedish population-based EIMS cohort, with clinical follow-up data from the Swedish MS Registry. Serum BA levels were analyzed using liquid chromatography-high-resolution mass spectrometry. Differential expression analysis was used to study differences in BAs between pwMS and controls. Cox proportional-hazard models were used to assess associations between BA concentrations and confirmed disability worsening (CDW) and the risk of reaching EDSS milestones 4.0 and 6.0.

**Results:** PwMS had lower concentrations of the primary conjugated BA, glycochenodeoxycholic acid (GCDCA, log_2_ FC -0.29, p=0.009) compared to controls. In relapsing-remitting MS compared to controls, lower concentrations of primary conjugated BAs (log2 FC -0.30, p=8.40E-5), secondary conjugated BAs (log2 FC -0.18, p=0.007), and total BAs (log2 FC -0.22, p=2.99E-4) were found. Sex-specific differences were also found, with male pwMS showing more substantial BA alterations. Elevated total BA levels were associated with increased risk for CDW (HR 1.22, 95% CI 1.08-1.39), driven mainly by primary conjugated (HR 1.19, 95% CI 1.06-1.33) and secondary conjugated BAs (HR 1.21, 95% CI 1.08-1.39).

**Conclusion:** This study identified alterations in serum BA profiles in pwMS compared to controls, with strong associations between conjugated BAs and the risk of disability worsening. These findings underscore the potential role of BAs in MS pathogenesis and disability worsening, suggesting they may be promising targets for future therapeutic interventions. Further research is warranted to clarify the underlying mechanisms of these associations.

## Background

Multiple sclerosis (MS) is a chronic, inflammatory, and demyelinating disease affecting the central nervous system (CNS). While the exact cause of MS remains largely unknown, it is believed to involve a combination of environmental factors and genetic susceptibility [1]. Environmental influences consistently linked to an increased risk of developing MS include smoking, adolescent obesity, and prior infection with the Epstein-Barr virus (EBV) [2]. However, factors influencing disease course have been studied less [3].

Changes in the gut microbiota have been observed in MS, with a reduction in beneficial bacteria and an increase in pro-inflammatory species [4,5]. These microbial alterations may contribute to systemic inflammation by disrupting the intestinal barrier and promoting immune activation. Gut dysbiosis in MS may also influence neuroinflammation and disease progression through the gut-brain axis, a communication network between the CNS and the gastrointestinal tract [4]. However, its importance for disease development and worsening remains to be determined. Bile acids (BAs), traditionally recognized for their function in digesting and absorbing dietary fats, also influence the colonic environment by modulating the composition and function of the gut microbiota. After synthesis in the liver and conjugation with taurine or glycine, BAs are stored in the gallbladder and released during digestion.

While most conjugated BAs are reabsorbed in the terminal ileum and recycled, a small fraction enters the colon, where they are metabolized by the microbiota into secondary bile acids. These microbial metabolites, in turn, influence gut homeostasis by modulating intestinal barrier integrity and shaping immune responses [6,7]. Conversely, BAs can regulate the gut microbiota directly through their antimicrobial activity or indirectly via interaction with nuclear and membrane receptors. This bidirectional interaction between BAs and the gut microbiota not only helps maintain intestinal barrier function and immune homeostasis but also provides resistance against opportunistic pathogens [8].

Bile acids are also essential messengers in the gut-brain axis communication [7]. They act through receptors like the farnesoid X receptor (FXR) and G-protein-coupled bile acid receptor 1 (GPBAR1 or TGR5), which regulate physiological processes, including immune responses, metabolism, and cellular homeostasis [7]. While BAs are primarily produced in the liver, recent findings show they can also be generated through alternative pathways in the brain and CNS [7]. Additionally, BAs can cross the blood-brain barrier and enter the CNS from the systemic circulation. Multiple BA receptors are present in the CNS, influencing immune responses and neuroinflammation [6,9–12]. In animal studies of EAE, supplementation with the BAs tauroursodeoxycholic acid (TUDCA) and taurochenodeoxycholic acid (TCDCA) resulted in lower clinical scores and ameliorated neuroinflammation [13,14]. The reduction in neuroinflammation was thought to be due to the activation of GPBAR1, leading to reduced astrocyte polarization. However, differences in BA metabolism and receptor function between murine models and humans pose challenges for translating these findings directly to human contexts [9].

The role of BAs in disease progression and, more specifically, disability worsening in relapsing-remitting MS (RRMS) and progressive MS (PMS) and in male and female people with MS (pwMS), respectively, has yet to be studied. Here, we investigated how serum BA levels in newly diagnosed pwMS were associated with confirmed disability worsening (CDW) as well as the probability of reaching the expanded disability status scale (EDSS) milestones 4.0 and 6.0.

## Methods

### Cohort

This study is based on the Swedish population-based case-control study, Epidemiological Investigation of Multiple Sclerosis (EIMS) [15]. Incident MS cases were recruited between 2005 and 2015 from neurological units across Sweden. All cases met the prevailing McDonald criteria [16,17]. For each case, matched controls (MC) were selected from the national population registry and matched by age, sex, and residential area. Participants completed detailed questionnaires on environmental exposures and lifestyle factors. MS cases were linked to the Swedish MS Registry (SMSreg) to obtain data on treatment, disability status (EDSS), and disability worsening [18]. The present study includes a subset of 907 pwMS and 907 MC with available questionnaire data, serum samples, and clinical follow-up data of the pwMS from the SMSreg.

### Bile acid analysis

Serum samples (50 µL) were thawed, and internal standards followed by ice-cold methanol were added for protein precipitation. The samples were centrifuged for 15 min at 21,100 RCF and 4°C and the sample supernatants were transferred to LC-vials and stored at -80°C until analysis. The internal standard mix contained glycocholic acid (GCA)-D4, glycoursodeoxycholic acid (GUDCA)-D4, taurocholic acid (TCA)-D4, and taurochenodeoxycholic acid (TCDCA)-D4. Quantitation was achieved through one-point calibration with the corresponding internal standard. A list of all targeted BAs, including abbreviations, can be found in *Supplementary Table S1*. Sample analysis was performed using a reversed-phase liquid chromatography column (Accucore C18, 100 × 2.1 mm, 2.6 μm, Thermo Scientific), with an Ultimate 3000 liquid chromatography system (Thermo Scientific) connected to a high-resolution hybrid quadrupole Q Exactive Orbitrap mass spectrometer (Thermo Scientific) operating in negative mode. The LC-HRMS method has been described in detail previously [19]. All data was processed using TraceFinder 4.1 (Thermo Scientific) and exported for further analysis.

### Statistical analysis

All computations were performed using R version 4.4.0 [20], and all concentrations were log_2_-transformed.

#### Correlation analysis

Spearman’s rank correlation analysis was performed to investigate the correlations between all BAs; the analysis was performed on all samples, as well as on pwMS and MC separately.

#### Differential expression analysis

Linear mixed-effect models were used for the differential expression analysis of the individual BAs, total BA concentration, and the sum of primary- and secondary non-conjugated, as well as primary- and secondary-conjugated BAs. The base model included covariates: age, sex, disease phenotype (MS, RRMS, and PMS), and body mass index (BMI, calculated as kg/m^2^). Additionally, lifestyle factors that could impact BA concentrations were included, and the ANOVA *P* values were assessed. These factors were regular smoking, irregular smoking, passive indoor smoking, Swedish snuff use, alcohol consumed (40%, cl) the week before sample collection, and treatment type. Treatments were categorized into first-line, second-line, and other therapies (*Supplementary Table S2*). Covariates with an ANOVA *P* value below 0.01 in any model were included in the final linear mixed-effect model. The final linear mixed-effect model included the following covariates: age, sex, disease phenotype, BMI, treatment, regular smoking, passive indoor smoking, and volume consumed alcohol (40%, cl). Interaction terms between sex and disease phenotype were also included in all models to explore potential sex differences. Four main clusters were found among the BAs in the correlation and hierarchical clustering analysis. There is, therefore, a strong dependence on measurements of BAs within each class. To account for multiple testing in the data analysis and the dependence of BA measurements within each of the four classes, the significance threshold was adjusted to p ≤ 0.0125.

#### Cox Proportional-Hazard model

Cox Proportional-Hazard models were used to evaluate if baseline BA concentrations affected time to CDW. Individuals without a registered EDSS in the SMSreg within three months of sample collection were excluded, leaving 551 pwMS. CDW was defined as follows: for EDSS of 0.0 to 1.0, the required increase was ≥1.5 points; for EDSS of 1.5 to 5.0, the required increase was ≥1.0 points; and for EDSS of ≥5.5, the required increase was ≥0.5 points [21,22]. Furthermore, the increasing EDSS needed to be sustained for at least three months to exclude temporary worsening due to relapses. The Cox Proportional-Hazard model included the following covariates: age, sex, BMI at the time of sample collection, regular smoking, ongoing treatment, disease phenotype, and baseline EDSS.

The same model assessed the effect of BA concentrations on the risk of reaching EDSS milestones 4.0 and 6.0, with the condition that these EDSS levels be maintained for at least three months. A p-value ≤ 0.0125 was considered significant after adjusting the significance level based on the four main clusters found in the correlation and hierarchical clustering analysis.

## Results

The analysis included 907 pwMS and 907 MC; of the 907 pwMS, 801 were diagnosed with RRMS, and 106 were diagnosed with PMS at the time of inclusion in the study. The study participants are described in *Table 1*. Clinical follow-up data, including EDSS scores, was collected from the SMSreg; the median follow-up time was 10.5±3.6 years. In total, 551 MS subjects had an EDSS score registered within three months from the time of inclusion and were included in the analyses regarding CDW and reaching EDSS milestones 4.0 and 6.0. There were no significant differences between the two cohorts. A description of the cohort subset included in the analysis of disability worsening can be found in *Supplementary Table S3*.

**Table 1.**
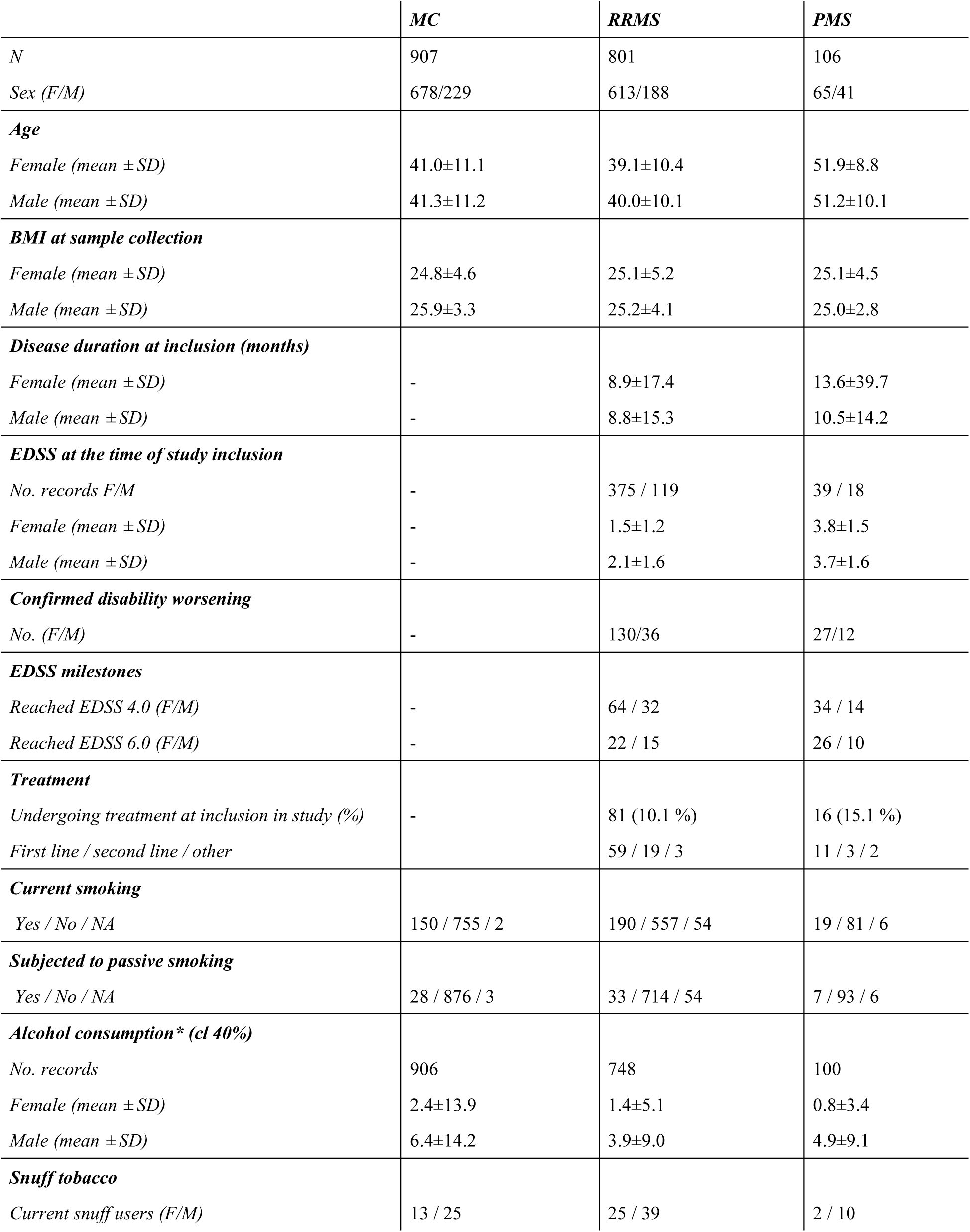
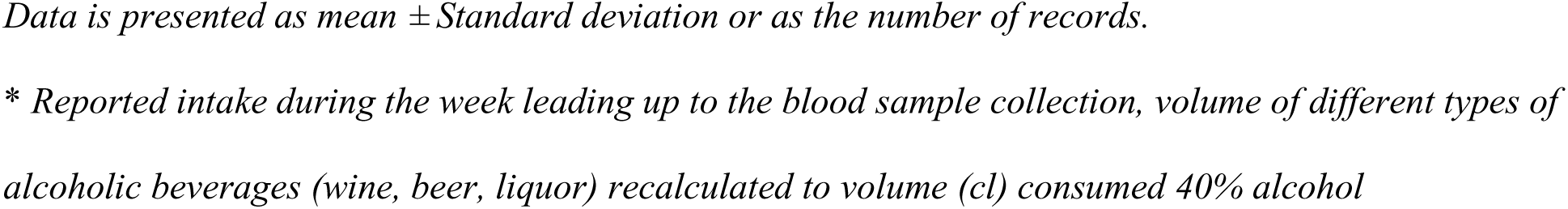
Selected characteristics of study participants from the EIMS cohort (n=1814).

|  | **MC** | **RRMS** | **PMS** |
| --- | --- | --- | --- |
| *N* | 907 | 801 | 106 |
| *Sex (F/M)* | 678/229 | 613/188 | 65/41 |
| **Age** |  |  |  |
| *Female (mean ± SD)* | 41.0±11.1 | 39.1±10.4 | 51.9±8.8 |
| *Male (mean ± SD)* | 41.3±11.2 | 40.0±10.1 | 51.2±10.1 |
| **BMI at sample collection** |  |  |  |
| *Female (mean ± SD)* | 24.8±4.6 | 25.1±5.2 | 25.1±4.5 |
| *Male (mean ± SD)* | 25.9±3.3 | 25.2±4.1 | 25.0±2.8 |
| **Disease duration at inclusion (months)** |  |  |  |
| *Female (mean ± SD)* | - | 8.9±17.4 | 13.6±39.7 |
| *Male (mean ± SD)* | - | 8.8±15.3 | 10.5±14.2 |
| **EDSS at the time of study inclusion** |  |  |  |
| *No. records F/M* | - | 375 / 119 | 39 / 18 |
| *Female (mean ± SD)* | - | 1.5±1.2 | 3.8±1.5 |
| *Male (mean ± SD)* | - | 2.1±1.6 | 3.7±1.6 |
| **Confirmed disability worsening** |  |  |  |
| *No. (F/M)* | - | 130/36 | 27/12 |
| **EDSS milestones** |  |  |  |
| *Reached EDSS 4.0 (F/M)* | - | 64 / 32 | 34 / 14 |
| *Reached EDSS 6.0 (F/M)* | - | 22 / 15 | 26 / 10 |
| **Treatment** |  |  |  |
| *Undergoing treatment at inclusion in study (%)* | - | 81 (10.1 %) | 16 (15.1 %) |
| *First line / second line / other* |  | 59 / 19 / 3 | 11 / 3 / 2 |
| **Current smoking** |  |  |  |
| *Yes / No / NA* | 150 / 755 / 2 | 190 / 557 / 54 | 19 / 81 / 6 |
| **Subjected to passive smoking** |  |  |  |
| *Yes / No / NA* | 28 / 876 / 3 | 33 / 714 / 54 | 7 / 93 / 6 |
| **Alcohol consumption\* (cl 40%)** |  |  |  |
| *No. records* | 906 | 748 | 100 |
| *Female (mean ± SD)* | 2.4±13.9 | 1.4±5.1 | 0.8±3.4 |
| *Male (mean ± SD)* | 6.4±14.2 | 3.9±9.0 | 4.9±9.1 |
| **Snuff tobacco** |  |  |  |
| *Current snuff users (F/M)* | 13 / 25 | 25 / 39 | 2 / 10 |

*Data is presented as mean ± Standard deviation or as the number of records.*
*\* Reported intake during the week leading up to the blood sample collection, volume of different types of alcoholic beverages (wine, beer, liquor) recalculated to volume (cl) consumed 40% alcohol*

### Bile acid metabolism is altered in MS

Strong correlations were observed among all bile acids, with a median correlation coefficient of r=0.473 and a median p-value of 3.189E−100 across all samples (pwMS and MC) and similar within the respective groups (pwMS: r=0.4745, p=7.16E−48, MC: r=0.4750, p=7.66E−51) (*Supplementary Figures S1–S3 and Tables S4–S6*). The high degree of correlation among BAs indicates their lack of independence. To assess the combined contributions of these BAs, we generated summed values for their respective classes: primary non-conjugated, primary conjugated, secondary non-conjugated, secondary conjugated, and total BAs. To account for multiple comparisons in the data analysis, we considered the four clusters (Supplementary Figure S1) as well as the four primary classes of BAs analyzed in the study as independent measures. Accordingly, we adjusted the significance threshold to p≤0.0125 for all analyses.

We performed differential expression analysis to investigate potential differences in serum BA concentration between pwMS and MC (*Figure 1A, Supplementary Table S7*). The model was adjusted for the following covariates: age, sex, disease phenotype, BMI, treatment, regular smoking, passive indoor smoking, and volume consumed alcohol (40%, cl). In MS, we generally found lower concentrations of BAs compared to MC. However, the only significant difference was the lower concentration of glycochenodeoxycholic acid (GCDCA) (log2 FC -0.291, p=0.009). When separating the analysis in RRMS and PMS groups, lower concentrations of primary conjugated BAs (log_2_ FC -0.301, p=8.40E-5), secondary conjugated BAs (log_2_ FC -0.184, p=0.007), and lower total BAs (log_2_ FC -0.220, p=2.99E-4) in RRMS were observed. In the PMS group, no significant changes were found.

**Figure 1.**
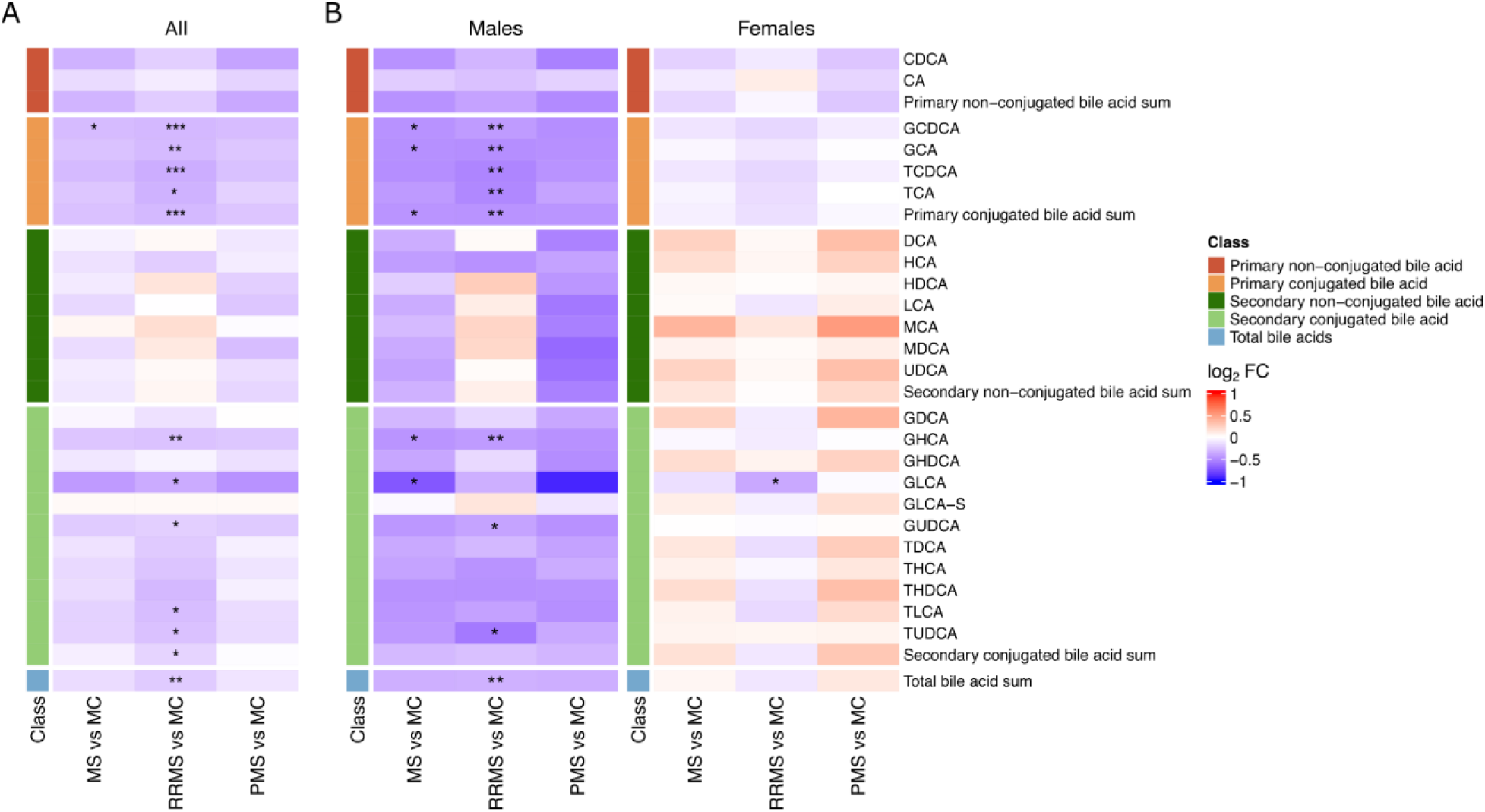
Heatmaps depicting the differences between MS, RRMS, PMS, and MC presented as log_2_ fold changes. (A) All pwMS and MC (B) stratified results based on sex. All individual foldchanges, including confidence intervals and p-values, are presented in supplementary Table S7-S9. The model included the following covariates: age, sex, disease phenotype, BMI, treatment, regular smoking, passive indoor smoking, and volume consumed alcohol (40%, cl). * P value≤0.0125; ** P value≤0.0025, *** P value≤2.5E-4.

Due to the large discrepancy in MS incidence between the sexes, we also stratified the analysis to investigate possible differences in BA metabolism between males and females (*Figure 1B, Supplementary Table S8&S9*). Significant differences in BA metabolism were observed between the sexes, with notable differences between male pwMS and MC, while the differences in female pwMS were less pronounced.

In men, we found lower concentrations, in the entire MS group and RRMS compared to MC, of total BAs (log_2_ FC -0.300, p=0.016, log_2_ FC -0.269, p=0.001) and primary conjugated BAs (log_2_ FC -0.424, p=0.009, log_2_ FC -0.474, p=0.4.25E-4). In stark contrast, in female pwMS with RRMS, lower concentrations were only found for GLCA (-0.374, p=0.002). No differences in BAs were found in women with PMS compared to MC.

### Bile acids are associated with confirmed disability worsening in MS

Using Cox-proportional hazard models adjusting for age, sex, BMI, regular smoking, ongoing treatment, disease phenotype, and baseline EDSS, we investigated the associations between the 24 different BA concentrations and CDW. A forest plot of all individually measured BAs and the summation of the respective BA classes and their association with CDW is presented in *Figure 2.* The sum of total BA was associated with an increased risk of CDW (HR 1.22, 95% CI 1.08 to 1.39). In more detail, this was mainly due to the associations of conjugated BAs with higher risks of CDW, both primary (HR 1.19, 95% CI 1.06 to 1.33) and secondary BAs (HR 1.21, 95% CI 1.08 to 1.39). The conjugated BAs significantly associated with CDW were GCDCA, GCA, TCDCA, TCA, GLCA-sulfate (GLCA-S), and taurodeoxycholic acid (TDCA).

**Figure 2.**
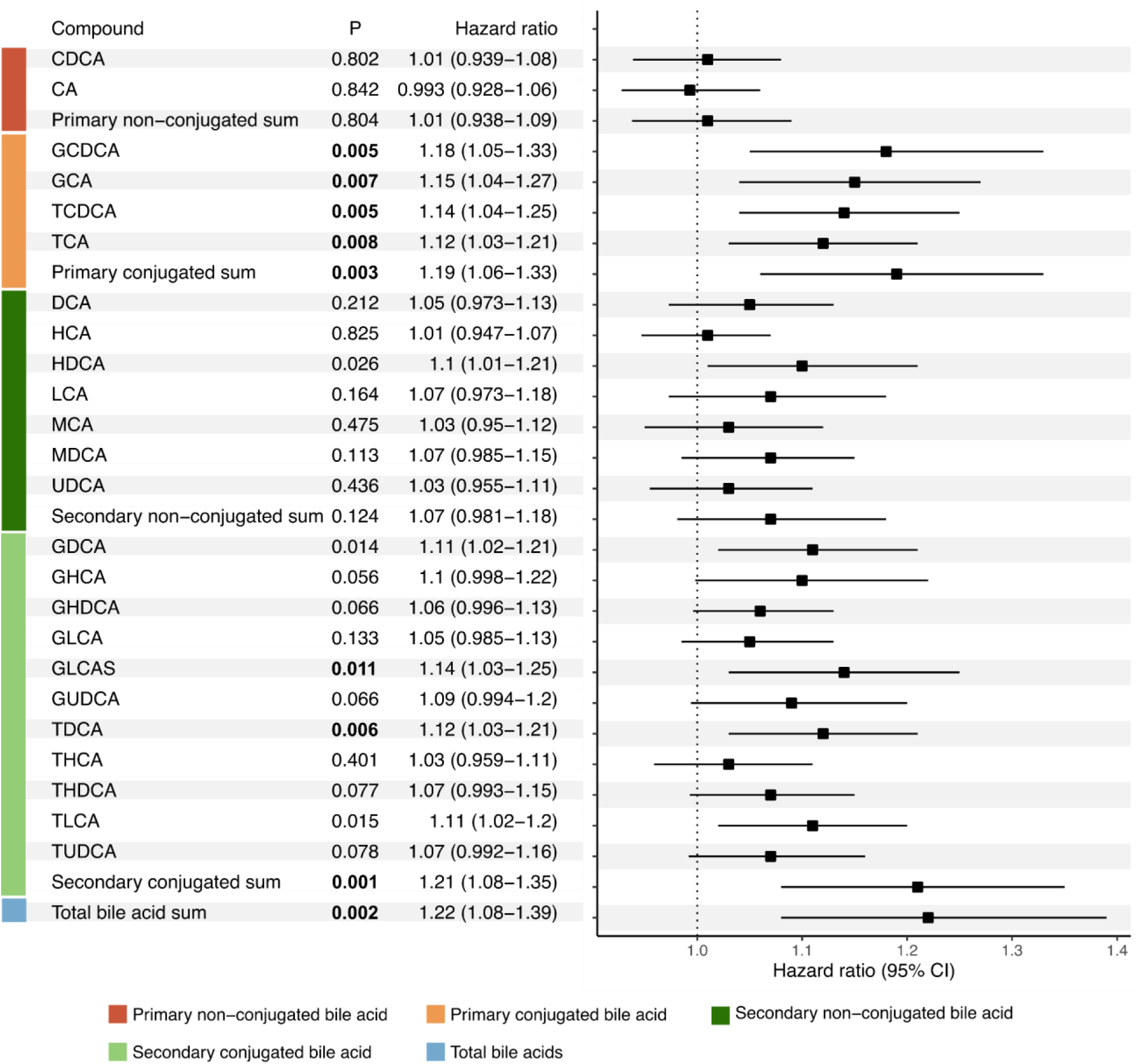
Hazard ratios, including 95% confidence intervals and p-values for bile acids and their association with confirmed disability worsening. Significant p-values (p≤0.0125) are highlighted with bold text. The model included the following covariates: age, sex, BMI, regular smoking, ongoing treatment, disease phenotype, and baseline EDSS.

Due to the significant differences in BA metabolism between male and female MS subjects in the differential expression analysis, we also stratified the analysis based on sex (*Table 2*). In male subjects, three conjugated BAs were found significantly associated with an increased risk of CDW, TCA (HR 1.22, 95% CI 1.05 to 1.43, p=0.011), GHDCA (HR 1.23, 95% CI 1.06 to 1.43, p=0.008) and TDCA (HR 1.22, 95% CI 1.05 to 1.43, p=0.012). In women, the sum of conjugated secondary BAs was significantly associated with an increased risk of CDW (HR 1.20, 95% CI 1.05 to 1.36, p=0.006).

**Table 2.**
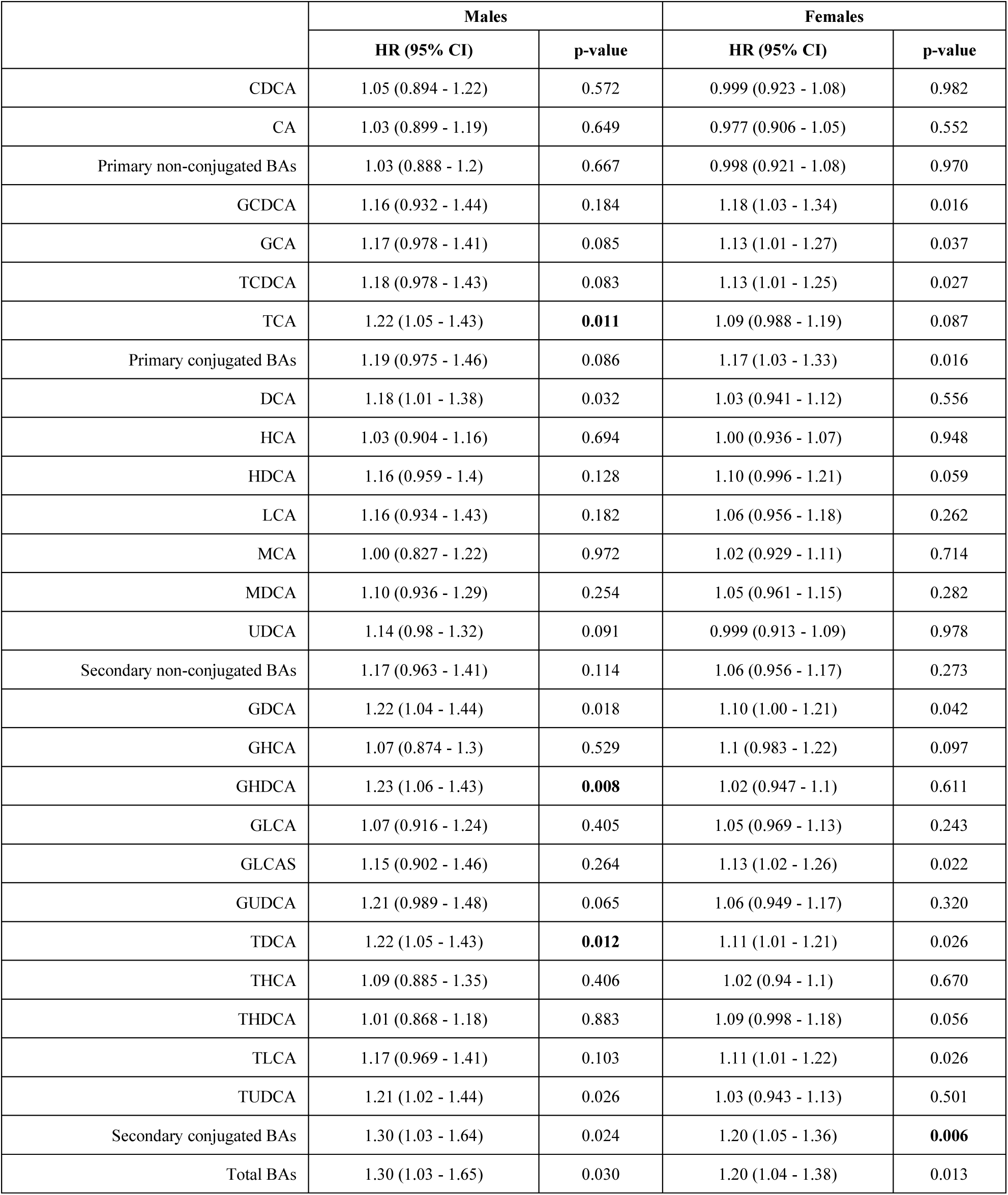
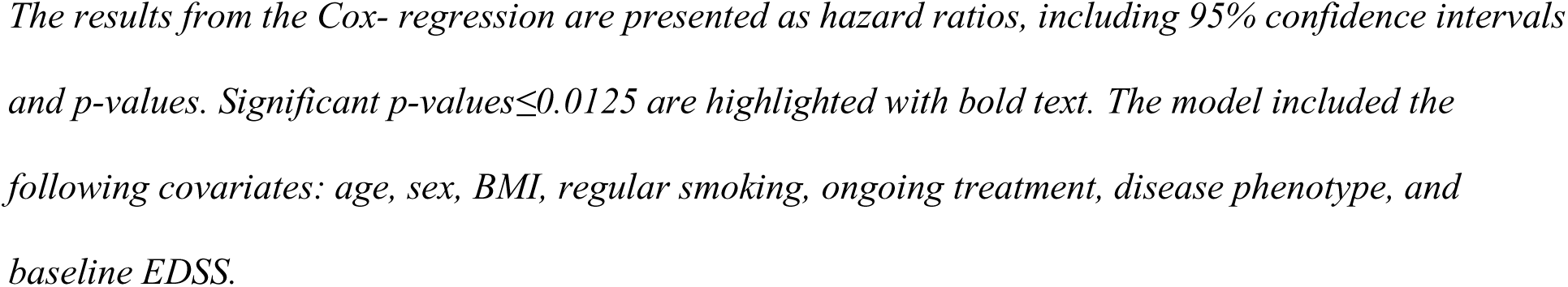
Cox regression of bile acids association with confirmed disability worsening (CDW) stratified based on sex.

|  | Males |  | Females |  |
| --- | --- | --- | --- | --- |
|  | HR (95% CI) | p-value | HR (95% CI) | p-value |
| CDCA | 1.05 (0.894 - 1.22) | 0.572 | 0.999 (0.923 - 1.08) | 0.982 |
| CA | 1.03 (0.899 - 1.19) | 0.649 | 0.977 (0.906 - 1.05) | 0.552 |
| Primary non-conjugated BAs | 1.03 (0.888 - 1.2) | 0.667 | 0.998 (0.921 - 1.08) | 0.970 |
| GCDCA | 1.16 (0.932 - 1.44) | 0.184 | 1.18 (1.03 - 1.34) | 0.016 |
| GCA | 1.17 (0.978 - 1.41) | 0.085 | 1.13 (1.01 - 1.27) | 0.037 |
| TCDCA | 1.18 (0.978 - 1.43) | 0.083 | 1.13 (1.01 - 1.25) | 0.027 |
| TCA | 1.22 (1.05 - 1.43) | **0.011** | 1.09 (0.988 - 1.19) | 0.087 |
| Primary conjugated BAs | 1.19 (0.975 - 1.46) | 0.086 | 1.17 (1.03 - 1.33) | 0.016 |
| DCA | 1.18 (1.01 - 1.38) | 0.032 | 1.03 (0.941 - 1.12) | 0.556 |
| HCA | 1.03 (0.904 - 1.16) | 0.694 | 1.00 (0.936 - 1.07) | 0.948 |
| HDCA | 1.16 (0.959 - 1.4) | 0.128 | 1.10 (0.996 - 1.21) | 0.059 |
| LCA | 1.16 (0.934 - 1.43) | 0.182 | 1.06 (0.956 - 1.18) | 0.262 |
| MCA | 1.00 (0.827 - 1.22) | 0.972 | 1.02 (0.929 - 1.11) | 0.714 |
| MDCA | 1.10 (0.936 - 1.29) | 0.254 | 1.05 (0.961 - 1.15) | 0.282 |
| UDCA | 1.14 (0.98 - 1.32) | 0.091 | 0.999 (0.913 - 1.09) | 0.978 |
| Secondary non-conjugated BAs | 1.17 (0.963 - 1.41) | 0.114 | 1.06 (0.956 - 1.17) | 0.273 |
| GDCA | 1.22 (1.04 - 1.44) | 0.018 | 1.10 (1.00 - 1.21) | 0.042 |
| GHCA | 1.07 (0.874 - 1.3) | 0.529 | 1.1 (0.983 - 1.22) | 0.097 |
| GHDCA | 1.23 (1.06 - 1.43) | **0.008** | 1.02 (0.947 - 1.1) | 0.611 |
| GLCA | 1.07 (0.916 - 1.24) | 0.405 | 1.05 (0.969 - 1.13) | 0.243 |
| GLCAS | 1.15 (0.902 - 1.46) | 0.264 | 1.13 (1.02 - 1.26) | 0.022 |
| GUDCA | 1.21 (0.989 - 1.48) | 0.065 | 1.06 (0.949 - 1.17) | 0.320 |
| TDCA | 1.22 (1.05 - 1.43) | **0.012** | 1.11 (1.01 - 1.21) | 0.026 |
| THCA | 1.09 (0.885 - 1.35) | 0.406 | 1.02 (0.94 - 1.1) | 0.670 |
| THDCA | 1.01 (0.868 - 1.18) | 0.883 | 1.09 (0.998 - 1.18) | 0.056 |
| TLCA | 1.17 (0.969 - 1.41) | 0.103 | 1.11 (1.01 - 1.22) | 0.026 |
| TUDCA | 1.21 (1.02 - 1.44) | 0.026 | 1.03 (0.943 - 1.13) | 0.501 |
| Secondary conjugated BAs | 1.30 (1.03 - 1.64) | 0.024 | 1.20 (1.05 - 1.36) | **0.006** |
| Total BAs | 1.30 (1.03 - 1.65) | 0.030 | 1.20 (1.04 - 1.38) | 0.013 |

*The results from the Cox- regression are presented as hazard ratios, including 95% confidence intervals and p-values. Significant p-values $\leq 0.0125$ are highlighted with bold text. The model included the following covariates: age, sex, BMI, regular smoking, ongoing treatment, disease phenotype, and baseline EDSS.*

### Bile acids are associated with the risk of reaching EDSS 4.0 in MS

Next, we investigated BA’s associations with the risk of reaching the disability milestones EDSS 4.0 and 6.0 (*Figure 3, Supplementary Table S10 and S11*). The Cox proportional hazards models were adjusted for age, sex, BMI, regular smoking, ongoing treatment, disease phenotype, and baseline EDSS. At inclusion, 31 pwMS had already reached EDSS 4.0, and 20 pwMS had reached EDSS 6.0, these pwMS were still included in the analysis. However, they were censored in the analysis as they were not at risk of reaching EDSS 4.0 or 6.0, respectively. After adjusting for multiple comparisons, the only significant association was for GDCA (HR 1.19, 95% CI 1.05 to 1.36, p=0.008). However, similar to the analysis of CDW we observed a trend towards an association with the risk of reaching EDSS 4.0 and total BAs (HR 1.17, 95% CI 1.02 to 1.34, p=0.025), as well as primary conjugated BAs (HR 1.16, 95% CI 1.03 to 1.30, p=0.018), and secondary conjugated BAs (HR 1.16, 95% CI 1.02 to 1.34, p=0.020).

**Figure 3.**
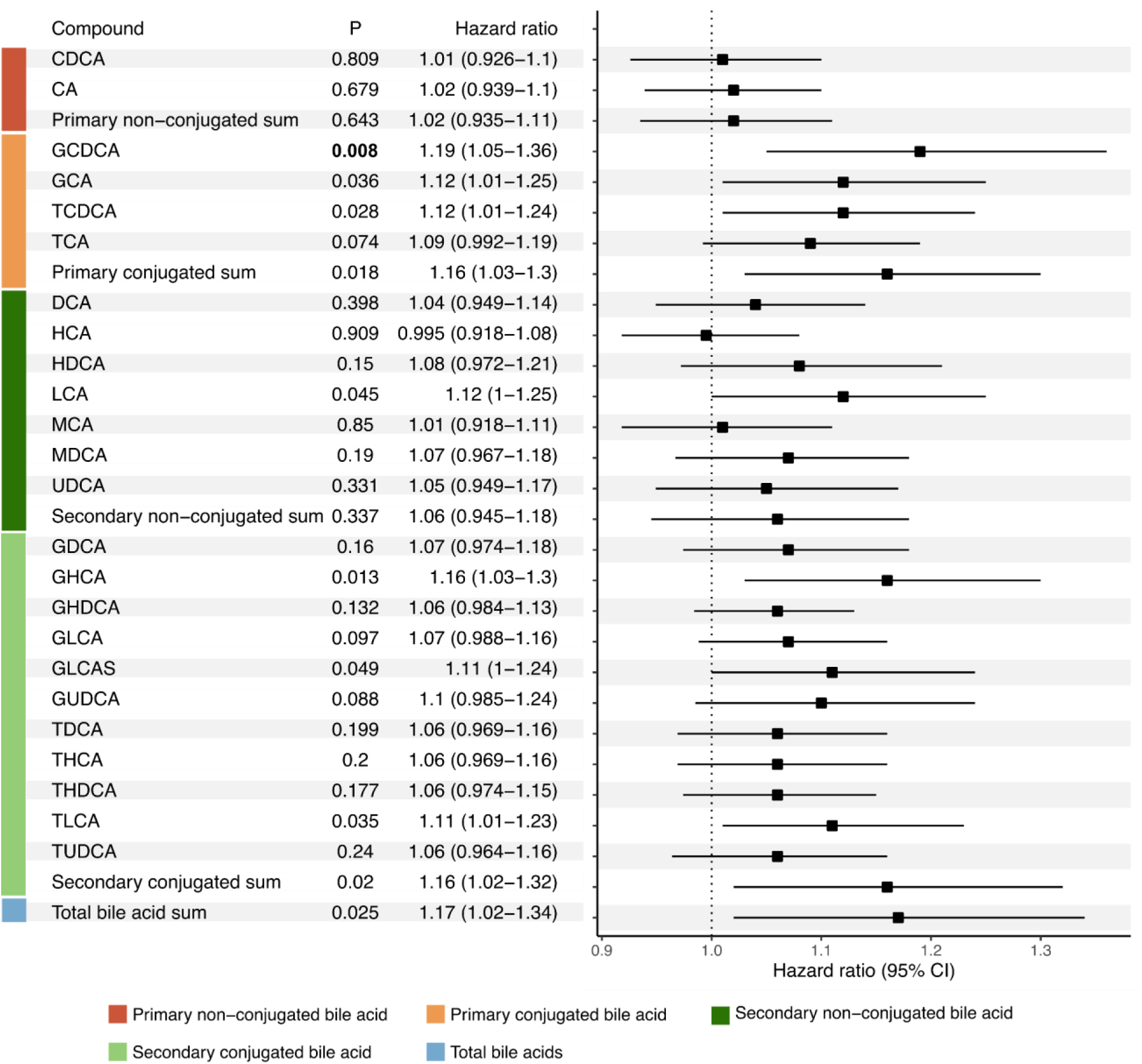
Hazard ratios, confidence intervals (95%), and p-values for bile acids and their association with risk of reaching EDSS 4.0. Significant p-values (p≤0.0125) are highlighted with bold text. The model included the following covariates: age, sex, BMI, regular smoking, ongoing treatment, disease phenotype, and baseline EDSS.

We performed the same analysis on male and female MS subjects (*Supplementary Table S10*). In female MS subjects, primary conjugated BAs (HR 1.24, 95% CI 1.05 to 1.48, p=0.012) and secondary conjugated BAs (HR 1.23, 95% CI 1.05 to 1.44, p=0.009) were associated with the risk of reaching EDSS 4.0. One individual conjugated BA was related to the risk of reaching EDSS 4.0, GCDCA (HR 1.26, 95% CI 1.06 to 1.50, p=0.008). In male MS subjects, the secondary conjugated BA glycohyodeoxycholic acid (GHDCA, HR 1.23, 95% CI 1.05 to 1.44, p=0.010) was associated with the risk of reaching EDSS 4.0.

Finally, we investigated the association of serum BA concentrations with the risk of reaching EDSS 6.0 but found no statistically significant associations (*Supplementary Table S11*). Due to the low number of events, especially in male MS subjects, no analysis stratified on sex was performed.

## Discussion

In this study, we found that serum BA profiles were different in pwMS compared to MC and that these differences were much more prominent in males than females. Furthermore, high concentrations of BAs were associated with the risk of disability worsening in MS. In particular, primary conjugated BAs were associated with an increased risk of CDW and the risk of reaching the EDSS milestone 4.0.

Differences in BA metabolism between pwMS and controls have been previously demonstrated in RRMS and PMS [13]. However, in contrast to our results, prior studies identified the most significant differences in the PMS group compared to controls. Whereas we found the most significant differences in the RRMS group. There are a few possible explanations for these different results between studies. In contrast to the previous study, most pwMS were treatment-naive when included in the current study. Furthermore, in this study, all comparions between MC and pwMS groups have been made with paired analyses, and pwMS and MC were matched on sex, age, and residential area. Serum BA concentrations have been reported to exhibit considerable variability across study populations and are influenced by factors such as age and sex [23,24]. This underscores the importance of matching pwMS and control subjects in the present study.

Consistent with earlier reports, we observed that most altered BAs were primary and secondary conjugated BAs [13]. Moreover, we observed significant sex-specific differences in BA metabolism. Male pwMS exhibited substantial differences in BA concentrations compared to MC, whereas such differences were minimal between female pwMS and their respective controls. This apparent sex difference in BA metabolism has not been reported in MS before. Similar sex differences in BA metabolism have been documented in Alzheimer’s disease (AD) [25,26]. A longitudinal study involving 1,180 participants (ranging from cognitively normal to those diagnosed with AD) found that changes in serum BA profiles were more pronounced, and occurred earlier, in male subjects compared to female subjects during disease development and progression [25]. The absence of longitudinal data on BA profiles prohibits us from conducting a similar analysis. However, given that the pwMS in our study were included relatively early in disease progression (mean 9.3 months from diagnosis), it is plausible that the sex differences we observe may be similar to those seen in AD.

Interestingly, we also found significant associations between serum BAs and increased risk of CDW and time to reach the milestone of EDSS 4.0. Even though total BA was linked to a higher risk of CDW, this association was primarily driven by primary conjugated BAs and, to some extent, secondary conjugated BAs. No significant association was observed with the time to reach EDSS 6.0. During the follow-up period, only 73 pwMS reached an EDSS score of 6.0, and the lack of statistical significance could be attributed to the relatively small number of pwMS reaching the outcome measure.

Blood biomarkers that can predict the risk of CDW or the risk of reaching specific EDSS scores are scarce in MS and have generally been limited to protein biomarkers. Neurofilament light chain (NfL) and glial fibrillary acidic protein (GFAP) are the most well-studied biomarkers that have been used to predict disability worsening [27–30]. Here, we observed that conjugated BAs measured early in the disease could possibly be used as biomarkers to predict the future risk of disability worsening in MS. However, while the mechanistic link between NfL and GFAP with disability worsening is relatively well understood, the mechanistic link between conjugated bile acids and disability worsening remains unclear.

In this study, we found that the concentrations of BAs were generally lower in pwMS as compared to controls. Yet, among pwMS, higher concentrations of conjugated BAs were associated with a higher risk of CDW. One possible explanation for these somewhat contrasting results could be that the BAs exert different effects at different stages in the disease progression. BAs interact with several receptors throughout the body, including FXR, GPBAR1, the vitamin D receptor (VDR), and the sphingosine-1-phosphate receptor 2 (S1PR-2). These interactions influence pro- and anti-inflammatory responses, immune function, and metabolism, which could impact disability worsening in MS. Some studies have reported that BAs act immunoregulatory by activating the FXR or GPBAR1 to reduce acute inflammation [9,31]. Lithocholic acid (LCA) and metabolites were found to reduce the differentiation of Th17 cells and increase Treg cell differentiation [32]. In contrast, others report that FXR antagonism or inactivation can promote the proliferation of regulatory T-cells [33]. Activation of GPBAR1 can change the polarization of macrophages from pro-inflammatory to anti-inflammatory and reduce astrocyte polarization [9,13,34]. However, other studies have investigated how conjugated BAs interact with S1PR2 to activate microglia and demonstrated that they contribute to neuroinflammation [10,11]. Activated microglia are often highlighted as one of the driving factors in sustained CNS inflammation and disease progression [35].

Hence BAs could possibly affect disease progression and disability worsening differently depending on the disease stage and possibly exert different effects in the long term and short term, respectively.

S1PRs are already targeted by disease-modifying treatments for MS, with S1PR modulators, such as fingolimod, primarily acting on S1PR-1 to block its effects [36]. Blocking the effects of S1PR-1 leads to inhibited cytokine amplification, reduced immune cell trafficking into the CNS, and reduced demyelination [36]. The conjugated BAs, found to be associated with CDW in this study, are known to bind to and activate S1PR-2 [37]. Activation of S1PR-2 leads to increased blood-brain barrier permeability, lymphatic endothelial cell permeability, and increased immune cell trafficking [38,39]. Activation of S1PR-2 can also contribute to microglial activation and pro-inflammatory polarization [10,11]. Moreover, S1PR-2 is over-expressed in the CNS of pwMS compared to healthy controls and more so in females than males [39]. This has been suggested to be one explanation for the susceptibility of CNS autoimmunity observed in females [39]. In an EAE model, treating mice with the S1PR-2 inhibitor JTE-013 resulted in reduced EAE disease severity, reduced BBB permeability, and CXCL-12 signaling [39]. Thus, the observed association between elevated serum concentrations of conjugated BAs and increased risk of disease progression may be attributed to the activation of S1PR-2 by conjugated BAs. Furthermore, S1PR-2 modulation could also be a potential target for future MS treatments.

Elevated serum concentrations of conjugated BAs may not directly contribute to the observed increased risk of disability worsening but could instead reflect other underlying factors associated with the risk of CDW. For example, inflammatory conditions, such as MS, are often linked to increased gut-barrier permeability, which may result in elevated serum BA levels [5]. Consequently, patients experiencing higher levels of inflammation might exhibit higher serum BA concentrations, potentially explaining the observed association between conjugated BAs and a higher risk of disability worsening [5,40].

Furthermore, the elevated concentrations of conjugated BA could result from a decreased capacity of the gut microbiota to deconjugate BAs, a process typically mediated by bile salt hydrolase (BSH) activity. Alterations in gut microbiota, which have been previously documented in MS, can lead to changes in BSH activity and, consequently, affect the BA pool [4,5]. However, the associations between specific alterations in gut microbiota and disease outcomes remain to be explored. Additionally, other factors that could alter the serum levels of BAs include less physical activity and slower bowel functions, which could be factors more common in pwMS at higher risk of disability worsening [41,42].

High levels of BAs (up to 100-fold higher compared to normal levels) are common in liver disease and are known to cause adverse neurological effects and disruption of the blood-brain barrier (BBB) [43]. Yet, even at concentrations lower than those typically seen in liver disease but similar to the levels we found in pwMS with the highest concentrations, BAs can still alter the BBB function, increasing its permeability [43]. Modulation of the BBB by BAs may also contribute to the observed association between BAs and disability worsening, as reported in this study.

In this large case-control study, we were able to show significant differences in BA serum profiles between pwMS and MC and its associations with disability worsening. However, some limitations of the study persist; we did not have information on the fasting state of the subjects at the time of sampling and were, therefore, unable to control for variations in serum BA levels during the day and in the fasting state. Additionally, we did not have information about the participant’s dietary patterns or caloric intake and could, therefore, not take this into account in the data analysis. Nevertheless, due to the large number of subjects in the analysis, it is unlikely that this has impacted the study outcome in a significant way. Moreover, we do not have longitudinal data regarding the BA levels and can, therefore, not draw any conclusions regarding possible changes in BA profiles and metabolism with disability worsening.

## Conclusions

In conclusion, our study provides evidence that altered bile acid profiles are associated with disability worsening in MS, highlighting the importance of bile acid metabolism in disease pathogenesis. These findings offer a promising foundation for future research into BA-modulating therapies as potential interventions for MS. Further studies are required to elucidate the precise mechanisms underlying these associations and to explore the therapeutic potential of targeting bile acid metabolism in slowing or preventing disability worsening in pwMS.

## Abbreviations

AD: Alzheimer’s disease
BA: Bile Acid
BBB: Blood-Brain Barrier
BMI: Body Mass Index
BSH: Bile Salt Hydrolase
CA: Cholic Acid
CDCA: Chenodeoxycholic Acid
CDW: Confirmed Disability Worsening
CNS: Central Nervous System
DCA: Deoxycholic Acid
EAE: Experimental Autoimmune Encephalomyelitis
EBV: Epstein-Barr Virus
EDSS: Expanded Disability Status Scale<colcnt=2>
EIMS: Epidemiological Investigation of Multiple Sclerosis
FXR: Farnesoid X Receptor
GPBAR1: G-Protein-coupled Bile Acid Receptor 1
GCA: Glycocholic Acid
GCDCA: Glycochenodeoxycholic Acid
GDCA: Glycodeoxycholic Acid
GFAP: Glial Fibrillary Acidic Protein
GHCA: Glycohyocholic Acid
GHDCA: Glycohyodeoxycholic Acid
GLCA: Glycolithocholic Acid
GLCA-S: Glycolithocholic Acid Sulfate
GUDCA: Glycoursodeoxycholic Acid
HCA: Hyocholic Acid
HDCA: Hyodeoxycholic Acid
LCA: Lithocholic Acid
MCA: Muricholic Acid
MDCA: Murideoxycholic Acid
MC: Matched Controls
MS: Multiple Sclerosis
NfL: Neurofilament Light Chain
PMS: Progressive Multiple Sclerosis
pwMS: people with Multiple Sclerosis
RRMS: Relapsing-Remitting Multiple Sclerosis
SMSreg: Swedish MS Registry
S1PR: Sphingosine-1-Phosphate Receptor
TCA: Taurocholic Acid
TCDCA: Taurochenodeoxycholic Acid
TDCA: Taurodeoxycholic Acid
THCA: Taurohyocholic Acid
TLCA: Taurolithocholic Acid
TUDCA: Tauroursodeoxycholic Acid
UDCA: Ursodeoxycholic Acid
VDR: Vitamin D Receptor

## Declarations

### Ethical statements

Sample collection and storage complied with local ethical standards and applicable national laws regarding human studies. The corresponding ethical approvals are EPN Uppsala: 2021-00702; EPN Stockholm: 04-252/1-4 and 2017/2386-32. All participants provided written informed consent.

### Consent for publication

Not applicable, as no identifiable human data or images are included in this manuscript.

### Data availability

Aggregated data supporting this study’s findings are available upon reasonable request from the corresponding author, KK. The data are not publicly available due to Swedish laws on personal integrity and health data, as well as the decision by the Ethics Committee.

### Author contributions

The EIMS cohort was conceptualized by IK, TO, AKH, and LA. The methods were developed by KK, HC, IE, and AV. AV and IE implemented the methods and analysis. The results were interpreted by KK, JB, IE, and AV. AV, IE, JB, and KK wrote the manuscript, and all the authors were involved in the review and editing process. All authors read and approved the final manuscript.

## Acknowledgment

We express our gratitude to all participants of the EIMS cohort. Additionally, we acknowledge the Swedish Neuro Registries/Multiple Sclerosis for the support.

