## Supplementary data for "Bile Acid Metabolism in Multiple Sclerosis is Perturbed and Associated with the Risk of Confirmed Disability Worsening"

Table S1 - Bile acid targets and calibration
Table S2 - Summary of treatments
Table S3 - Description of general and clinical characteristics for the 551 PwMS included in the analysis of disability worsening
Figure S1 - Correlation heatmap including r-values for all samples (pwMS and MC)
Figure S2 - Correlation heatmap including r-values for pwMS samples
Figure S3 - Correlation heatmap including r-values for MC samples
Table S4 - P-values from correlation analysis, for all samples (pwMS and MC)
Table S5 - P-values from correlation analysis, for pwMS group
Table S6 - P-values from correlation analysis, for MC group
Table S7 - Heat map data for Figure 1A, foldchanges (95% CI) and p-values.
Table S8 - Heat map data for Figure 1B males, foldchanges (95% CI) and p-values.
Table S9 - Heat map data for Figure 1B females, foldchanges (95% CI) and p-values.
Table S10 - EDSS 4 males and females, cox regression results
Table S11 - EDSS 6 all MS, cox regression results

*Supplementary Table S1. Summary of bile acid targets, m/z, retention time, and internal standard used for calibration.*

| **Bile acid** | **Abbreviation** | **m/z** | **RT** | **IS** |
|---|---|---|---|---|
| *Chenodeoxycholic acid* | CDCA | 391.28555 | 8.74 | TCDCA-D4 |
| *Cholic acid* | CA | 407.28049 | 8.16 | TCDCA-D4 |
| *Glycochenodeoxycholic acid* | GCDCA | 448.30652 | 7.9 | TCDCA-D4 |
| *Glycocholic acid* | GCA | 464.30148 | 7.37 | GCA-D4 |
| *Taurochenodeoxycholic acid* | TCDCA | 498.28949 | 7.8 | TCDCA-D4 |
| *Taurocholic acid* | TCA | 514.28412 | 7.25 | TCA-D4 |
| *Deoxycholic acid* | DCA | 391.28555 | 8.86 | TCDCA-D4 |
| *Hyocholic acid* | HCA | 407.28049 | 7.88 | TCDCA-D4 |
| *Hyodeoxycholic acid* | HDCA | 391.28555 | 8.16 | TCDCA-D4 |
| *Lithocholic acid* | LCA | 375.290525 | 9.33 | TCDCA-D4 |
| *Muricholic acid* | MCA | 407.28049 | 7.43 | GCA-D4 |
| *Murideoxycholic acid* | MDCA | 391.28555 | 7.69 | TCDCA-D4 |
| *Ursodeoxycholic acid* | UDCA | 391.28555 | 7.88 | TCDCA-D4 |
| *Glycodeoxycholic acid* | GDCA | 448.30652 | 8.08 | TCDCA-D4 |
| *Glycohyocholic acid* | GHCA | 464.30148 | 6.91 | GUDCA-D4 |
| *Glycohyodeoxycholic acid* | GHCDA | 448.30652 | 7.2 | TCA-D4 |
| *Glycolithocholic acid* | GLCA | 432.31202 | 8.53 | TCDCA-D4 |
| *Glycolithocholic acid 3-sulfate* | GLCA-S | 512.26855 | 7.5 | GCA-D4 |
| *Glycoursodeoxycholic acid* | GUDCA | 448.30652 | 6.94 | GUDCA-D4 |
| *Taurodeoxycholic acid* | TDCA | 498.28949 | 7.96 | TCDCA-D4 |
| *Taurohyocholic acid* | THCA | 514.28412 | 6.8 | GUDCA-D4 |
| *Taurohyodeoxycholic acid* | THDCA | 498.28949 | 7.05 | GUDCA-D4 |
| *Taurolithocholic acid* | TLCA | 482.29453 | 8.4 | TCDCA-D4 |
| *Tauroursodeoxycholic acid* | TUDCA | 498.28949 | 6.83 | GUDCA-D4 |
| *Glycocholic acid-D4* | GCA-D4 | 468.32687 | 7.37 | |
| *Glycoursodeoxycholic acid-D4* | GUDCA-D4 | 452.33195 | 6.94 | |
| *Taurochenodeoxycholic acid-D4* | TCDCA-D4 | 502.31459 | 7.8 | |
| *Taurocholic acid-D4* | TCA-D4 | 518.3095 | 7.25 | |

*Supplementary Table S2. Treatments included in the different treatment groups*

| **Treatment category** | **Treatment** |
|---|---|
| First line DMT | *Teriflunomid, Interferon beta-1a, Interferon beta-1b, Glatiramer, Dimethylfumarat, Intravenous immunoglobulin (IVIG), Peginterferon beta-1a,* |
| Second line DMT | *Fingolimod, HSCT/BEAM, HSCT/CYK, Ofatumumab, Alemtuzumab, Kladribin, Mitoxantron, Okrelizumab, Rituximab, Natalizumab, Daklizumab beta* |
| Other treatments | *Erenumab, Prednison, Metylprednisolon* |

*Supplementary Table S3*. General and clinical characteristics for the 551 individuals included in the Cox proportional hazard analysis regarding confirmed disability worsening (CDW).

| | *RRMS* | *PMS* |
|---|---|---|
| *N* | 494 | 57 |
| *Sex (F/M)* | 375 / 119 | 39 / 18 |
| ***Age*** | | |
| *Female (mean ± SD)* | 38.9±10.3 | 51.9±8.7 |
| *Male (mean ± SD)* | 37.3±10.1 | 48.9±8.2 |
| ***BMI at sample collection*** | | |
| *Female (mean ± SD)* | 25.1±5.2 | 25.4±4.7 |
| *Male (mean ± SD)* | 25.2±4.3 | 25.7±2.8 |
| ***Disease duration at inclusiom (months)*** | | |
| *Female (mean ± SD)* | 9.5±20.5 | 9.6±11.5 |
| *Male (mean ± SD)* | 9.3±15.1 | 10.5±10.3 |
| ***Number of hospital visits*** | | |
| *Female (mean ± SD)* | 11.5±5.6 | 8.9±4.7 |
| *Male (mean ± SD)* | 12.3±6.7 | 8.9±4.6 |
| ***Follow-up time (years)*** | | |
| *Female (mean ± SD)* | 10.7±3.5 | 9.0±4.1 |
| *Male (mean ± SD)* | 10.2±3.7 | 9.0±4.1 |
| ***EDSS at the time of study inclusion*** | | |
| *Female (mean ± SD)* | 1.5±1.2 | 3.8±1.5 |
| *Male (mean ± SD)* | 2.1±1.6 | 3.7±1.6 |
| ***EDSS milestones*** | | |
| *Reached EDSS 4 witin study follow-up (F/M)* | 64 / 32 | 34 / 14 |
| *Reached EDSS 6 witin study follow-up (F/M)* | 22 / 15 | 26 / 10 |
| ***Treatment*** | | |
| *Undergoing treatment at inclusion in study (%)* | 8.3 % | 15.8 % |
| *First line, second line, other* | 25,13,3 | 5,3,1 |
| ***Current smoking*** | | |
| *No. Yes/No/NA* | 118 / 338 / 38 | 9 / 44 / 4 |
| ***Subjected to passive smoking*** | | |
| *No. Yes/No/NA* | 15 / 440 / 39 | 3 / 50 / 4 |
| ***Alcohol consumption* (cl 40%)*** | | |
| *No. records* | 457 | 53 |
| *Female (mean ± SD)* | 0.9±2.9 | 0.6±1.6 |
| *Male (mean ± SD)* | 4.2±9.6 | 3.6±5.7 |
| ***Snuff tobacco*** | | |
| *Current snuff users (F/M)* | 16 / 23 | 0 / 5 |

* *Reported intake the week leading up to blood sample collection, amount of different types of alcoholic beverages (wine, beer, liquor) recalculated to volume (cl) consumed 40% alcohol*

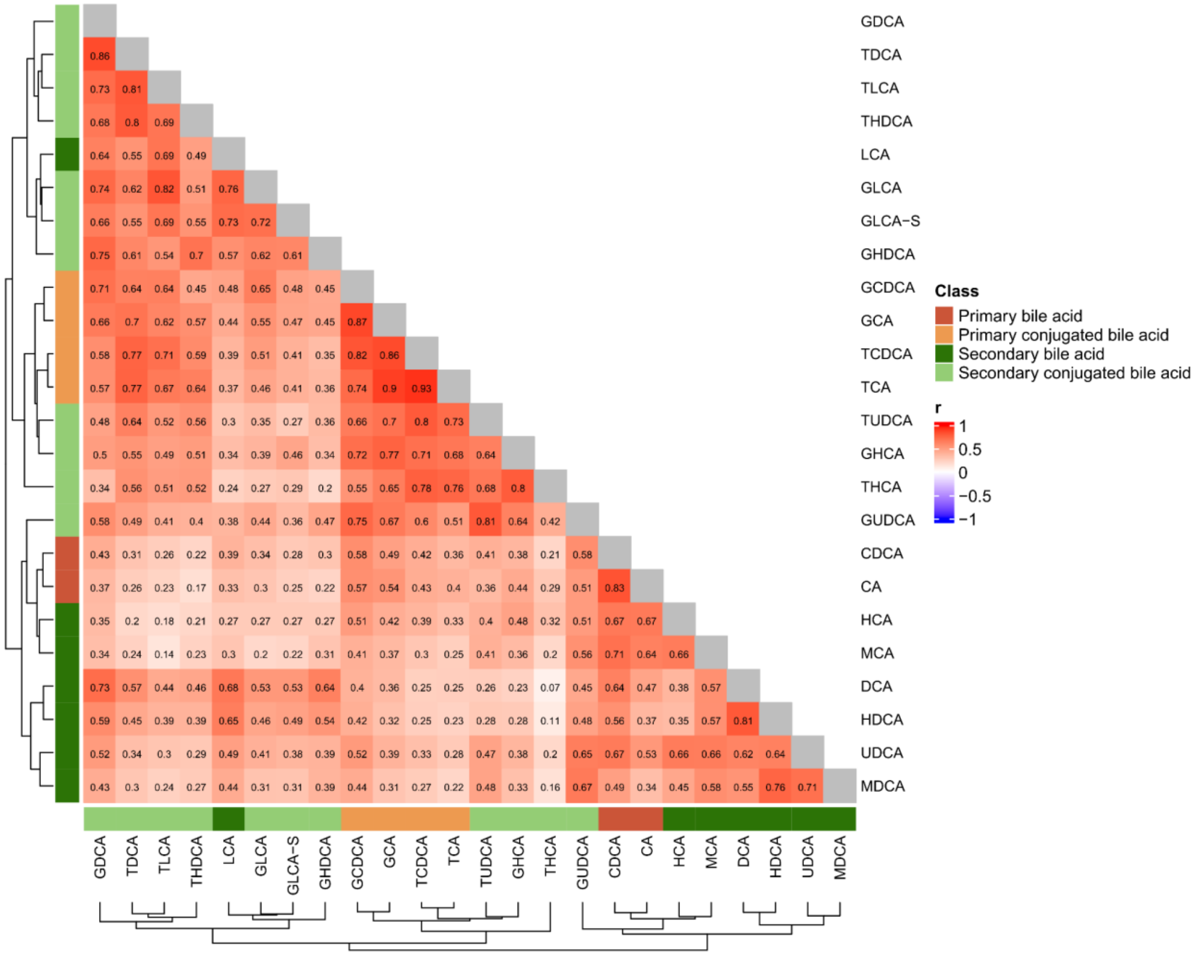


*Supplementary Figure S1. Correlation heat map for all BAs, including r values. The correlations are based on all samples, pwMS and MC.*

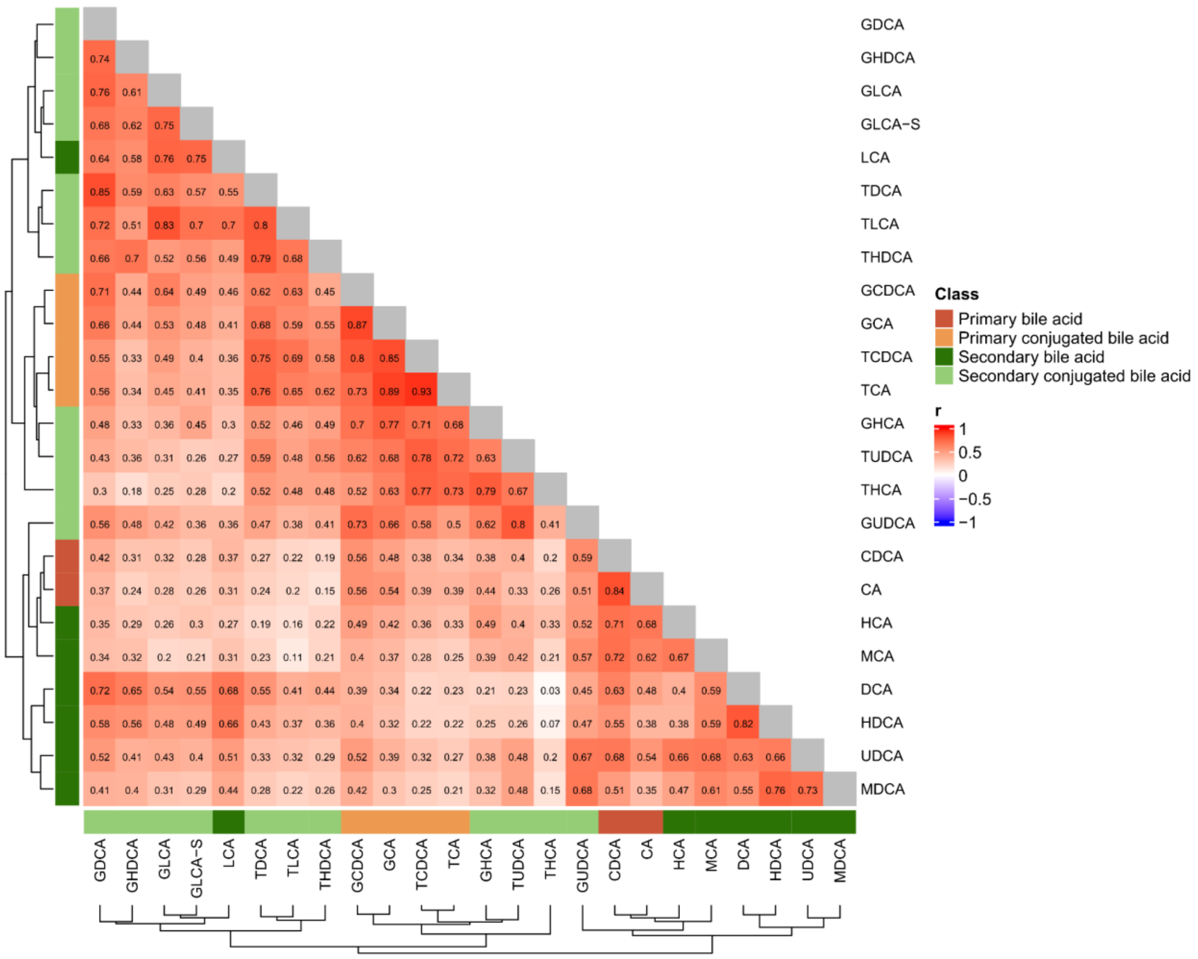


*Supplementary Figure S2. Correlation heat map for all BAs, including r values. The correlations are based on the pwMS group.*

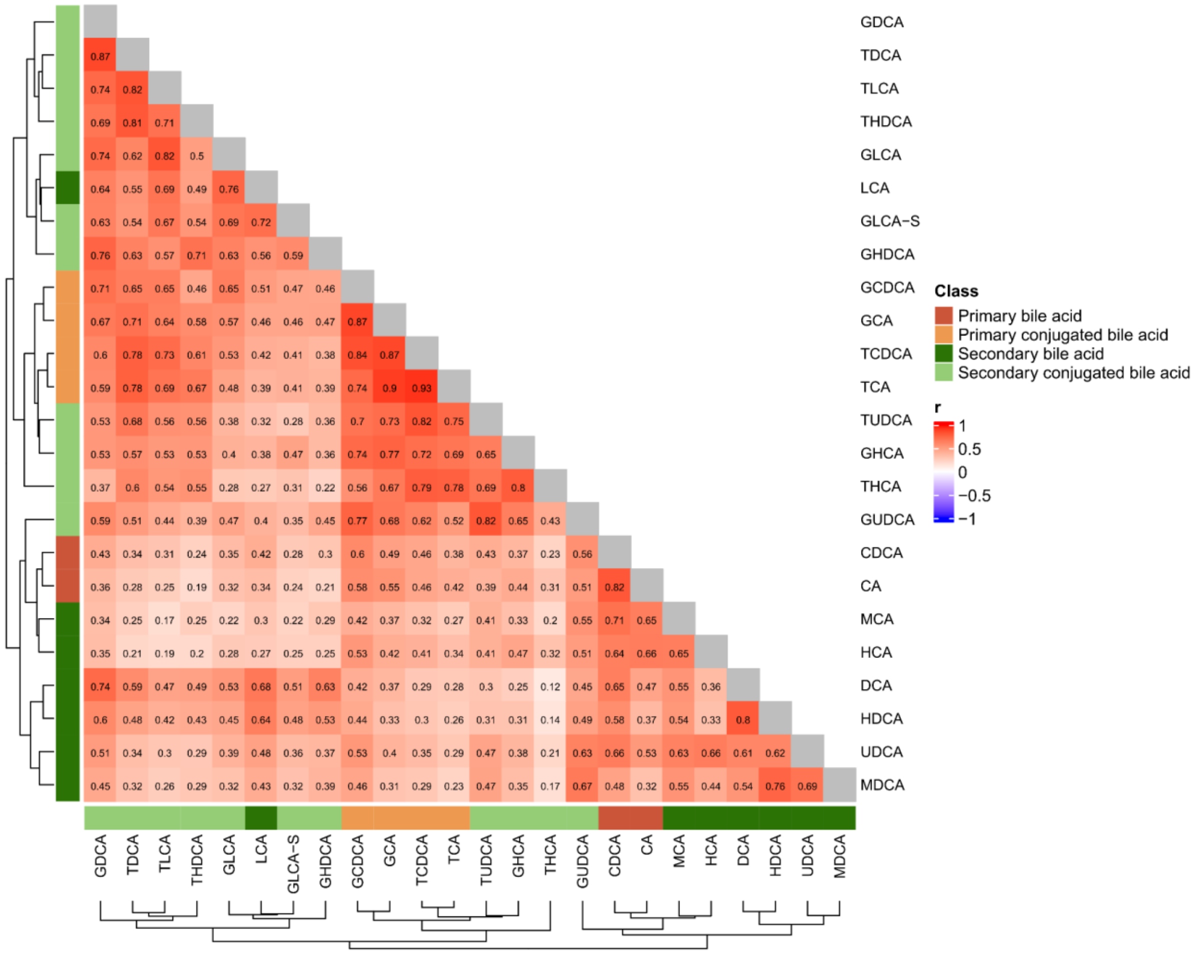


*Supplementary Figure S3. Correlation heat map for all BAs, including r values. The correlations are based on the MC group.*

*Supplementary Table S4*. p-values from correlation analyses (Spearman) between BAs for all samples, pwMS and MC.

| | *CDCA* | *CA* | *GCDCA* | *GCA* | *TCDCA* | *TCA* | *DCA* | *HCA* | *HDCA* | *LCA* | *MCA* | *MDCA* | *UDCA* | *GDCA* | *GHCA* | *GHDCA* | *GLCA* | *GLCA-S* | *GUDCA* | *TDCA* | *THCA* | *THDCA* | *TLCA* | *TUDCA* |
|---|---|---|---|---|---|---|---|---|---|---|---|---|---|---|---|---|---|---|---|---|---|---|---|---|
| *CDCA* | NA | 0 | 3.6E-162 | 1.2E-109 | 2.1E-79 | 1.5E-57 | 1.8E-206 | 1.3E-239 | 7.5E-151 | 3.5E-67 | 6.4E-243 | 3.8E-112 | 1.0E-231 | 1.3E-80 | 8.2E-63 | 2.9E-39 | 7.9E-49 | 9.6E-34 | 9.7E-160 | 2.0E-40 | 3.9E-18 | 1.0E-18 | 1.6E-29 | 4.9E-74 |
| *CA* | 0 | NA | 3.6E-157 | 5.2E-140 | 4.7E-81 | 5.0E-72 | 1.2E-100 | 2.2E-234 | 1.4E-60 | 8.7E-46 | 9.4E-183 | 5.8E-50 | 2.6E-132 | 8.2E-59 | 4.6E-86 | 1.5E-21 | 6.3E-39 | 3.8E-27 | 9.2E-123 | 9.0E-30 | 1.7E-32 | 5.2E-12 | 1.8E-22 | 7.6E-56 |
| *GCDCA* | 3.6E-162 | 3.6E-157 | NA | 0 | 0 | 1.2E-310 | 5.8E-71 | 4.6E-120 | 5.1E-77 | 7.1E-104 | 4.3E-65 | 7.0E-85 | 3.9E-127 | 1.1E-279 | 7.5E-290 | 5.9E-89 | 3.0E-214 | 7.5E-106 | 0.0 | 4.6E-208 | 8.0E-129 | 1.2E-83 | 1.6E-210 | 9.3E-227 |
| *GCA* | 1.2E-109 | 5.2E-140 | 0 | NA | 0 | 0 | 1.3E-55 | 1.4E-79 | 9.9E-46 | 2.3E-83 | 1.8E-51 | 1.1E-40 | 1.9E-68 | 1.2E-231 | 0 | 5.7E-91 | 1.6E-145 | 2.3E-100 | 7.0E-238 | 1.9E-264 | 1.2E-200 | 6.0E-141 | 5.0E-190 | 4.3E-268 |
| *TCDCA* | 2.1E-79 | 4.7E-81 | 0 | 0 | NA | 0 | 7.5E-27 | 6.3E-65 | 3.8E-28 | 2.2E-64 | 2.3E-33 | 2.0E-31 | 6.6E-48 | 2.2E-161 | 7.0E-282 | 3.8E-52 | 3.7E-121 | 4.8E-73 | 1.8E-179 | 0 | 0 | 1.0E-154 | 4.3E-280 | 0 |
| *TCA* | 1.5E-57 | 5.0E-72 | 1.2E-310 | 0 | 0 | NA | 5.3E-28 | 9.5E-48 | 4.2E-24 | 5.2E-59 | 9.9E-25 | 2.0E-20 | 2.3E-33 | 1.7E-159 | 1.0E-249 | 4.3E-57 | 1.2E-96 | 6.8E-73 | 5.8E-119 | 0 | 3.9E-304 | 8.1E-191 | 4.4E-237 | 6.1E-302 |
| *DCA* | 1.8E-206 | 1.2E-100 | 5.8E-71 | 1.3E-55 | 7.5E-27 | 5.3E-28 | NA | 4.9E-63 | 0 | 1.0E-240 | 2.4E-136 | 5.4E-143 | 3.4E-196 | 6.6E-299 | 1.1E-23 | 3.1E-206 | 9.3E-132 | 3.8E-131 | 9.7E-91 | 2.6E-155 | 3.2E-03 | 8.6E-86 | 1.5E-86 | 1.3E-29 |
| *HCA* | 1.3E-239 | 2.2E-234 | 4.6E-120 | 1.4E-79 | 6.3E-65 | 9.5E-48 | 4.9E-63 | NA | 1.9E-54 | 3.0E-31 | 3.4E-201 | 7.2E-93 | 5.5E-227 | 5.3E-54 | 1.8E-104 | 2.2E-31 | 7.4E-32 | 1.7E-32 | 3.7E-122 | 2.7E-18 | 3.9E-41 | 1.9E-17 | 4.2E-14 | 5.0E-71 |
| *HDCA* | 7.5E-151 | 1.4E-60 | 5.1E-77 | 9.9E-46 | 3.8E-28 | 4.2E-24 | 0 | 1.9E-54 | NA | 1.0E-216 | 2.7E-135 | 0 | 5.0E-208 | 1.1E-169 | 2.5E-33 | 1.4E-135 | 3.5E-96 | 9.0E-108 | 6.3E-105 | 5.1E-92 | 1.6E-05 | 4.7E-61 | 2.3E-67 | 1.2E-34 |
| *LCA* | 3.5E-67 | 8.7E-46 | 7.1E-104 | 2.3E-83 | 2.2E-64 | 5.2E-59 | 1.0E-240 | 3.0E-31 | 1.0E-216 | NA | 2.5E-34 | 2.6E-83 | 1.9E-110 | 1.5E-204 | 8.2E-50 | 6.8E-150 | 0 | 2.0E-300 | 3.9E-62 | 2.9E-141 | 4.9E-22 | 3.2E-100 | 4.2E-255 | 7.8E-38 |
| *MCA* | 6.4E-243 | 9.4E-183 | 4.3E-65 | 1.8E-51 | 2.3E-33 | 9.9E-25 | 2.4E-136 | 3.4E-201 | 2.7E-135 | 2.5E-34 | NA | 8.3E-143 | 6.2E-197 | 1.6E-43 | 5.0E-49 | 1.5E-35 | 4.4E-16 | 1.4E-18 | 2.6E-133 | 6.5E-22 | 5.8E-15 | 3.4E-19 | 3.6E-08 | 4.2E-65 |
| *MDCA* | 3.8E-112 | 5.8E-50 | 7.0E-85 | 1.1E-40 | 2.0E-31 | 2.0E-20 | 5.4E-143 | 7.2E-93 | 0 | 2.6E-83 | 8.3E-143 | NA | 1.0E-279 | 6.4E-82 | 9.6E-47 | 1.7E-66 | 7.5E-42 | 6.3E-41 | 1.7E-238 | 5.2E-38 | 1.3E-10 | 4.5E-29 | 3.8E-25 | 7.5E-102 |
| *UDCA* | 1.0E-231 | 2.6E-132 | 3.9E-127 | 1.9E-68 | 6.6E-48 | 2.3E-33 | 3.4E-196 | 5.5E-227 | 5.0E-208 | 1.9E-110 | 6.2E-197 | 1.0E-279 | NA | 2.5E-124 | 2.8E-63 | 9.3E-67 | 3.0E-73 | 1.0E-63 | 3.4E-220 | 1.7E-49 | 7.5E-17 | 3.4E-32 | 5.3E-40 | 2.1E-101 |
| *GDCA* | 1.3E-80 | 8.2E-59 | 1.1E-279 | 1.2E-231 | 2.2E-161 | 1.7E-159 | 6.6E-299 | 5.3E-54 | 1.1E-169 | 1.5E-204 | 1.6E-43 | 6.4E-82 | 2.5E-124 | NA | 8.4E-117 | 9.6E-321 | 3.0e-319 | 5.2E-225 | 3.8E-163 | 0 | 8.8E-45 | 3.0E-218 | 1.1E-298 | 8.8E-105 |
| *GHCA* | 8.2E-63 | 4.6E-86 | 7.5E-290 | 0 | 7.0E-282 | 1.0E-249 | 1.1E-23 | 1.8E-104 | 2.5E-33 | 8.2E-50 | 5.0E-49 | 9.6E-47 | 2.8E-63 | 8.4E-117 | NA | 5.5E-50 | 1.5E-65 | 3.6E-97 | 1.1E-205 | 1.2E-140 | 0 | 7.3E-109 | 2.4E-111 | 1.6E-207 |
| *GHDCA* | 2.9E-39 | 1.5E-21 | 5.9E-89 | 5.7E-91 | 3.8E-52 | 4.3E-57 | 3.1E-206 | 2.2E-31 | 1.4E-135 | 6.8E-150 | 1.5E-35 | 1.7E-66 | 9.3E-67 | 9.6E-321 | 5.5E-50 | NA | 6.7E-189 | 6.3E-181 | 8.4E-97 | 4.9E-182 | 6.7E-16 | 4.3E-239 | 3.6E-134 | 6.0E-54 |
| *GLCA* | 7.9E-49 | 6.3E-39 | 3.0E-214 | 1.6E-145 | 3.7E-121 | 1.2E-96 | 9.3E-132 | 7.4E-32 | 3.5E-96 | 0 | 4.4E-16 | 7.5E-42 | 3.0E-73 | 3.0e-319 | 1.5E-65 | 6.7E-189 | NA | 8.3E-283 | 9.4E-88 | 9.8E-196 | 7.9E-29 | 2.5E-107 | 0 | 2.6E-52 |
| *GLCA-S* | 9.6E-34 | 3.8E-27 | 7.5E-106 | 2.3E-100 | 4.8E-73 | 6.8E-73 | 3.8E-131 | 1.7E-32 | 9.0E-108 | 2.0E-300 | 1.4E-18 | 6.3E-41 | 1.0E-63 | 5.2E-225 | 3.6E-97 | 6.3E-181 | 8.3E-283 | NA | 1.1E-55 | 7.1E-147 | 6.4E-34 | 1.4E-128 | 6.3E-254 | 7.5E-32 |
| *GUDCA* | 9.7E-160 | 9.2E-123 | 0 | 7.0E-238 | 1.8E-179 | 5.8E-119 | 9.7E-91 | 3.7E-122 | 6.3E-105 | 3.9E-62 | 2.6E-133 | 1.7E-238 | 3.4E-220 | 3.8E-163 | 1.1E-205 | 8.4E-97 | 9.4E-88 | 1.1E-55 | NA | 7.9E-110 | 1.8E-71 | 6.6E-63 | 1.4E-73 | 0 |
| *TDCA* | 2.0E-40 | 9.0E-30 | 4.6E-208 | 1.9E-264 | 0 | 0 | 2.6E-155 | 2.7E-18 | 5.1E-92 | 2.9E-141 | 6.5E-22 | 5.2E-38 | 1.7E-49 | 0 | 1.2E-140 | 4.9E-182 | 9.8E-196 | 7.1E-147 | 7.9E-110 | NA | 6.2E-139 | 0 | 0 | 5.4E-205 |
| *THCA* | 3.9E-18 | 1.7E-32 | 8.0E-129 | 1.2E-200 | 0 | 3.9E-304 | 3.2E-03 | 3.9E-41 | 1.6E-05 | 4.9E-22 | 5.8E-15 | 1.3E-10 | 7.5E-17 | 8.8E-45 | 0 | 6.7E-16 | 7.9E-29 | 6.4E-34 | 1.8E-71 | 6.2E-139 | NA | 1.7E-103 | 1.8E-107 | 1.7E-221 |
| *THDCA* | 1.0E-18 | 5.2E-12 | 1.2E-83 | 6.0E-141 | 1.0E-154 | 8.1E-191 | 8.6E-86 | 1.9E-17 | 4.7E-61 | 3.2E-100 | 3.4E-19 | 4.5E-29 | 3.4E-32 | 3.0E-218 | 7.3E-109 | 4.3E-239 | 2.5E-107 | 1.4E-128 | 6.6E-63 | 0 | 1.7E-103 | NA | 5.2E-232 | 6.2E-135 |
| *TLCA* | 1.6E-29 | 1.8E-22 | 1.6E-210 | 5.0E-190 | 4.3E-280 | 4.4E-237 | 1.5E-86 | 4.2E-14 | 2.3E-67 | 4.2E-255 | 3.6E-08 | 3.8E-25 | 5.3E-40 | 1.1E-298 | 2.4E-111 | 3.6E-134 | 0 | 6.3E-254 | 1.4E-73 | 0 | 1.8E-107 | 5.2E-232 | NA | 9.8E-127 |
| *TUDCA* | 4.9E-74 | 7.6E-56 | 9.3E-227 | 4.3E-268 | 0 | 6.1E-302 | 1.3E-29 | 5.0E-71 | 1.2E-34 | 7.8E-38 | 4.2E-65 | 7.5E-102 | 2.1E-101 | 8.8E-105 | 1.6E-207 | 6.0E-54 | 2.6E-52 | 7.5E-32 | 0 | 5.4E-205 | 1.7E-221 | 6.2E-135 | 9.8E-127 | NA |

*Supplementary Table S5*. p-values from correlation analyses (Spearman) between BAs for pwMS group.

| | *CDCA* | *CA* | *GCDCA* | *GCA* | *TCDCA* | *TCA* | *DCA* | *HCA* | *HDCA* | *LCA* | *MCA* | *MDCA* | *UDCA* | *GDCA* | *GHCA* | *GHDCA* | *GLCA* | *GLCA-S* | *GUDCA* | *TDCA* | *THCA* | *THDCA* | *TLCA* | *TUDCA* |
|---|---|---|---|---|---|---|---|---|---|---|---|---|---|---|---|---|---|---|---|---|---|---|---|---|
| *CDCA* | NA | 0 | 3.6E-162 | 1.2E-109 | 2.1E-79 | 1.5E-57 | 1.8E-206 | 1.3E-239 | 7.5E-151 | 3.5E-67 | 6.4E-243 | 3.8E-112 | 1.0E-231 | 1.3E-80 | 8.2E-63 | 2.9E-39 | 7.9E-49 | 9.6E-34 | 9.7E-160 | 2.0E-40 | 3.9E-18 | 1.0E-18 | 1.6E-29 | 4.9E-74 |
| *CA* | 0 | NA | 3.6E-157 | 5.2E-140 | 4.7E-81 | 5.0E-72 | 1.2E-100 | 2.2E-234 | 1.4E-60 | 8.7E-46 | 9.4E-183 | 5.8E-50 | 2.6E-132 | 8.2E-59 | 4.6E-86 | 1.5E-21 | 6.3E-39 | 3.8E-27 | 9.2E-123 | 9.0E-30 | 1.7E-32 | 5.2E-12 | 1.8E-22 | 7.6E-56 |
| *GCDCA* | 3.6E-162 | 3.6E-157 | NA | 0 | 0 | 1.23E-310 | 5.8E-71 | 4.6E-120 | 5.1E-77 | 7.1E-104 | 4.3E-65 | 7.0E-85 | 3.9E-127 | 1.1E-279 | 7.5E-290 | 5.9E-89 | 3.0E-214 | 7.5E-106 | 0 | 4.6E-208 | 8.0E-129 | 1.2E-83 | 1.6E-210 | 9.3E-227 |
| *GCA* | 1.2E-109 | 5.2E-140 | 0 | NA | 0 | 0 | 1.3E-55 | 1.4E-79 | 9.9E-46 | 2.3E-83 | 1.8E-51 | 1.1E-40 | 1.9E-68 | 1.2E-231 | 0 | 5.7E-91 | 1.6E-145 | 2.3E-100 | 7.0E-238 | 1.9E-264 | 1.2E-200 | 6.0E-141 | 5.0E-190 | 4.3E-268 |
| *TCDCA* | 2.1E-79 | 4.7E-81 | 0 | 0 | NA | 0 | 7.5E-27 | 6.3E-65 | 3.8E-28 | 2.2E-64 | 2.3E-33 | 2.0E-31 | 6.6E-48 | 2.2E-161 | 7.0E-282 | 3.8E-52 | 3.7E-121 | 4.8E-73 | 1.8E-179 | 0 | 0 | 1.0E-154 | 4.3E-280 | 0 |
| *TCA* | 1.5E-57 | 5.0E-72 | 1.2E-310 | 0 | 0 | NA | 5.3E-28 | 9.5E-48 | 4.2E-24 | 5.2E-59 | 9.9E-25 | 2.0E-20 | 2.3E-33 | 1.7E-159 | 1.0E-249 | 4.3E-57 | 1.2E-96 | 6.8E-73 | 5.8E-119 | 0 | 3.9E-304 | 8.1E-191 | 4.4E-237 | 6.1E-302 |
| *DCA* | 1.8E-206 | 1.2E-100 | 5.8E-71 | 1.3E-55 | 7.5E-27 | 5.3E-28 | NA | 4.9E-63 | 0 | 1.0E-240 | 2.4E-136 | 5.4E-143 | 3.4E-196 | 6.6E-299 | 1.1E-23 | 3.1E-206 | 9.3E-132 | 3.8E-131 | 9.7E-91 | 2.6E-155 | 3.2E-03 | 8.6E-86 | 1.5E-86 | 1.3E-29 |
| *HCA* | 1.3E-239 | 2.2E-234 | 4.6E-120 | 1.4E-79 | 6.3E-65 | 9.5E-48 | 4.9E-63 | NA | 1.9E-54 | 3.0E-31 | 3.4E-201 | 7.2E-93 | 5.5E-227 | 5.3E-54 | 1.8E-104 | 2.2E-31 | 7.4E-32 | 1.7E-32 | 3.7E-122 | 2.7E-18 | 3.9E-41 | 1.9E-17 | 4.2E-14 | 5.0E-71 |
| *HDCA* | 7.5E-151 | 1.4E-60 | 5.1E-77 | 9.9E-46 | 3.8E-28 | 4.2E-24 | 0 | 1.9E-54 | NA | 1.0E-216 | 2.7E-135 | 0 | 5.0E-208 | 1.1E-169 | 2.5E-33 | 1.4E-135 | 3.5E-96 | 9.0E-108 | 6.3E-105 | 5.1E-92 | 1.6E-05 | 4.7E-61 | 2.3E-67 | 1.2E-34 |
| *LCA* | 3.5E-67 | 8.7E-46 | 7.1E-104 | 2.3E-83 | 2.2E-64 | 5.2E-59 | 1.0E-240 | 3.0E-31 | 1.0E-216 | NA | 2.5E-34 | 2.6E-83 | 1.9E-110 | 1.5E-204 | 8.2E-50 | 6.8E-150 | 0 | 2.0E-300 | 3.9E-62 | 2.9E-141 | 4.9E-22 | 3.2E-100 | 4.2E-255 | 7.8E-38 |
| *MCA* | 6.4E-243 | 9.4E-183 | 4.3E-65 | 1.8E-51 | 2.3E-33 | 9.9E-25 | 2.4E-136 | 3.4E-201 | 2.7E-135 | 2.5E-34 | NA | 8.3E-143 | 6.2E-197 | 1.6E-43 | 5.0E-49 | 1.5E-35 | 4.4E-16 | 1.4E-18 | 2.6E-133 | 6.5E-22 | 5.8E-15 | 3.4E-19 | 3.6E-08 | 4.2E-65 |
| *MDCA* | 3.8E-112 | 5.8E-50 | 7.0E-85 | 1.1E-40 | 2.0E-31 | 2.0E-20 | 5.4E-143 | 7.2E-93 | 0 | 2.6E-83 | 8.3E-143 | NA | 1.0E-279 | 6.4E-82 | 9.6E-47 | 1.7E-66 | 7.5E-42 | 6.3E-41 | 1.7E-238 | 5.2E-38 | 1.3E-10 | 4.5E-29 | 3.8E-25 | 7.5E-102 |
| *UDCA* | 1.0E-231 | 2.6E-132 | 3.9E-127 | 1.9E-68 | 6.6E-48 | 2.3E-33 | 3.4E-196 | 5.5E-227 | 5.0E-208 | 1.9E-110 | 6.2E-197 | 1.0E-279 | NA | 2.5E-124 | 2.8E-63 | 9.3E-67 | 3.0E-73 | 1.0E-63 | 3.4E-220 | 1.7E-49 | 7.5E-17 | 3.4E-32 | 5.3E-40 | 2.1E-101 |
| *GDCA* | 1.3E-80 | 8.2E-59 | 1.1E-279 | 1.2E-231 | 2.2E-161 | 1.7E-159 | 6.6E-299 | 5.3E-54 | 1.1E-169 | 1.5E-204 | 1.6E-43 | 6.4E-82 | 2.5E-124 | NA | 8.4E-117 | 9.6E-321 | 3.0e-319 | 5.2E-225 | 3.8E-163 | 0 | 8.8E-45 | 3.0E-218 | 1.1E-298 | 8.8E-105 |
| *GHCA* | 8.2E-63 | 4.6E-86 | 7.5E-290 | 0 | 7.0E-282 | 1.0E-249 | 1.1E-23 | 1.8E-104 | 2.5E-33 | 8.2E-50 | 5.0E-49 | 9.6E-47 | 2.8E-63 | 8.4E-117 | NA | 5.5E-50 | 1.5E-65 | 3.6E-97 | 1.1E-205 | 1.2E-140 | 0 | 7.3E-109 | 2.4E-111 | 1.6E-207 |
| *GHDCA* | 2.9E-39 | 1.5E-21 | 5.9E-89 | 5.7E-91 | 3.8E-52 | 4.3E-57 | 3.1E-206 | 2.2E-31 | 1.4E-135 | 6.8E-150 | 1.5E-35 | 1.7E-66 | 9.3E-67 | 9.6E-321 | 5.5E-50 | NA | 6.7E-189 | 6.3E-181 | 8.4E-97 | 4.9E-182 | 6.7E-16 | 4.3E-239 | 3.6E-134 | 6.0E-54 |
| *GLCA* | 7.9E-49 | 6.3E-39 | 3.0E-214 | 1.6E-145 | 3.7E-121 | 1.2E-96 | 9.3E-132 | 7.4E-32 | 3.5E-96 | 0 | 4.4E-16 | 7.5E-42 | 3.0E-73 | 3.0e-319 | 1.5E-65 | 6.7E-189 | NA | 8.3E-283 | 9.4E-88 | 9.8E-196 | 7.9E-29 | 2.5E-107 | 0 | 2.6E-52 |
| *GLCA-S* | 9.6E-34 | 3.8E-27 | 7.5E-106 | 2.3E-100 | 4.8E-73 | 6.8E-73 | 3.8E-131 | 1.7E-32 | 9.0E-108 | 2.0E-300 | 1.4E-18 | 6.3E-41 | 1.0E-63 | 5.2E-225 | 3.6E-97 | 6.3E-181 | 8.3E-283 | NA | 1.1E-55 | 7.1E-147 | 6.4E-34 | 1.4E-128 | 6.3E-254 | 7.5E-32 |
| *GUDCA* | 9.7E-160 | 9.2E-123 | 0 | 7.0E-238 | 1.8E-179 | 5.8E-119 | 9.7E-91 | 3.7E-122 | 6.3E-105 | 3.9E-62 | 2.6E-133 | 1.7E-238 | 3.4E-220 | 3.8E-163 | 1.1E-205 | 8.4E-97 | 9.4E-88 | 1.1E-55 | NA | 7.9E-110 | 1.8E-71 | 6.6E-63 | 1.4E-73 | 0 |
| *TDCA* | 2.0E-40 | 9.0E-30 | 4.6E-208 | 1.9E-264 | 0 | 0 | 2.6E-155 | 2.7E-18 | 5.1E-92 | 2.9E-141 | 6.5E-22 | 5.2E-38 | 1.7E-49 | 0 | 1.2E-140 | 4.9E-182 | 9.8E-196 | 7.1E-147 | 7.9E-110 | NA | 6.2E-139 | 0 | 0 | 5.4E-205 |
| *THCA* | 3.9E-18 | 1.7E-32 | 8.0E-129 | 1.2E-200 | 0 | 3.9E-304 | 3.2E-03 | 3.9E-41 | 1.6E-05 | 4.9E-22 | 5.8E-15 | 1.3E-10 | 7.5E-17 | 8.8E-45 | 0 | 6.7E-16 | 7.9E-29 | 6.4E-34 | 1.8E-71 | 6.2E-139 | NA | 1.7E-103 | 1.8E-107 | 1.7E-221 |
| *THDCA* | 1.0E-18 | 5.2E-12 | 1.2E-83 | 6.0E-141 | 1.0E-154 | 8.1E-191 | 8.6E-86 | 1.9E-17 | 4.7E-61 | 3.2E-100 | 3.4E-19 | 4.5E-29 | 3.4E-32 | 3.0E-218 | 7.3E-109 | 4.3E-239 | 2.5E-107 | 1.4E-128 | 6.6E-63 | 0 | 1.7E-103 | NA | 5.2E-232 | 6.2E-135 |
| *TLCA* | 1.6E-29 | 1.8E-22 | 1.6E-210 | 5.0E-190 | 4.3E-280 | 4.4E-237 | 1.5E-86 | 4.2E-14 | 2.3E-67 | 4.2E-255 | 3.6E-08 | 3.8E-25 | 5.3E-40 | 1.1E-298 | 2.4E-111 | 3.6E-134 | 0 | 6.3E-254 | 1.4E-73 | 0 | 1.8E-107 | 5.2E-232 | NA | 9.8E-127 |
| *TUDCA* | 4.9E-74 | 7.6E-56 | 9.3E-227 | 4.3E-268 | 0 | 6.1E-302 | 1.3E-29 | 5.0E-71 | 1.2E-34 | 7.8E-38 | 4.2E-65 | 7.5E-102 | 2.1E-101 | 8.8E-105 | 1.6E-207 | 6.0E-54 | 2.6E-52 | 7.5E-32 | 0 | 5.4E-205 | 1.7E-221 | 6.2E-135 | 9.8E-127 | NA |

*Supplementary Table S6*. p-values from correlation analyses (Spearman) between BAs for MC group.

| | *CDCA* | *CA* | *GCDCA* | *GCA* | *TCDCA* | *TCA* | *DCA* | *HCA* | *HDCA* | *LCA* | *MCA* | *MDCA* | *UDCA* | *GDCA* | *GHCA* | *GHDCA* | *GLCA* | *GLCA-S* | *GUDCA* | *TDCA* | *THCA* | *THDCA* | *TLCA* | *TUDCA* |
|---|---|---|---|---|---|---|---|---|---|---|---|---|---|---|---|---|---|---|---|---|---|---|---|---|
| *CDCA* | NA | 1.5E-216 | 2.46E-88 | 5.77E-57 | 3.21E-48 | 2.41E-32 | 2.1E-109 | 1.6E-106 | 2.97E-82 | 1.39E-39 | 3.7E-122 | 5.06E-53 | 1.8E-112 | 1.83E-42 | 3.89E-31 | 6.15E-20 | 8.24E-28 | 5.39E-18 | 2.57E-77 | 1.49E-25 | 5.6E-11 | 5.21E-12 | 7.83E-21 | 6.65E-41 |
| *CA* | 1.5E-216 | NA | 2.05E-83 | 8.41E-73 | 3.72E-48 | 8.45E-40 | 1.98E-50 | 5E-113 | 2.48E-30 | 6.54E-26 | 6.09E-98 | 1.06E-23 | 1.52E-65 | 2.2E-29 | 1.83E-43 | 3.92E-10 | 1.14E-22 | 2.01E-13 | 3.07E-62 | 7.39E-18 | 1.49E-19 | 8.92E-08 | 1.69E-14 | 2.0E-33 |
| *GCDCA* | 2.46E-88 | 2.05E-83 | NA | 2.1E-284 | 2.7E-241 | 3E-159 | 7.77E-40 | 3.79E-66 | 1.54E-44 | 9.96E-60 | 3.1E-36 | 3.43E-49 | 7.07E-68 | 4.4E-141 | 8.8E-156 | 1.1E-47 | 8E-110 | 2.94E-50 | 5.7E-176 | 5.9E-110 | 4.11E-71 | 3.83E-43 | 9.3E-110 | 2.1E-131 |
| *GCA* | 5.77E-57 | 8.41E-73 | 2.1E-284 | NA | 1.5E-277 | 0 | 3.52E-31 | 7.46E-41 | 3.56E-25 | 6.26E-49 | 8.86E-28 | 4.96E-22 | 1.73E-35 | 1.3E-118 | 2.1E-176 | 2.16E-50 | 8.11E-78 | 1.93E-49 | 5.6E-122 | 1.8E-139 | 1.2E-109 | 2.95E-75 | 1E-104 | 4.6E-149 |
| *TCDCA* | 3.21E-48 | 3.72E-48 | 2.7E-241 | 1.5E-277 | NA | 0 | 1.97E-18 | 2.02E-37 | 5.79E-20 | 5.31E-39 | 4.89E-21 | 1.4E-19 | 3.16E-28 | 3.72E-91 | 5.6E-146 | 2.32E-31 | 6.12E-66 | 2.92E-38 | 2.51E-98 | 5.5E-187 | 5.1E-179 | 3.26E-83 | 4.4E-151 | 1.4E-223 |
| *TCA* | 2.41E-32 | 8.45E-40 | 3E-159 | 0 | 0 | NA | 1.48E-17 | 2.29E-25 | 2.66E-15 | 2.21E-33 | 1.11E-14 | 6.17E-12 | 8.86E-19 | 3.68E-86 | 7.2E-127 | 1.2E-33 | 2.54E-52 | 1.45E-37 | 6.21E-63 | 3E-185 | 3.1E-168 | 2.7E-104 | 3.5E-129 | 7E-163 |
| *DCA* | 2.1E-109 | 1.98E-50 | 7.77E-40 | 3.52E-31 | 1.97E-18 | 1.48E-17 | NA | 2.55E-28 | 1.8E-205 | 1.7E-121 | 1.68E-63 | 2.64E-70 | 5.88E-95 | 1.6E-155 | 8.66E-15 | 1.1E-100 | 4.82E-67 | 3.32E-60 | 1.72E-47 | 1.89E-85 | 0.00082 | 4.15E-49 | 2.01E-51 | 9.64E-20 |
| *HCA* | 1.6E-106 | 5E-113 | 3.79E-66 | 7.46E-41 | 2.02E-37 | 2.29E-25 | 2.55E-28 | NA | 4.91E-24 | 7.3E-17 | 2.15E-98 | 5.07E-43 | 1.8E-113 | 2.01E-27 | 1.02E-49 | 3.46E-14 | 1.08E-17 | 5.77E-14 | 1.48E-60 | 1.62E-10 | 5.05E-21 | 1.21E-08 | 6.06E-09 | 3.87E-37 |
| *HDCA* | 2.97E-82 | 2.48E-30 | 1.54E-44 | 3.56E-25 | 5.79E-20 | 2.66E-15 | 1.8E-205 | 4.91E-24 | NA | 2.1E-105 | 1.35E-60 | 3.4E-172 | 6.87E-96 | 1.63E-88 | 1.54E-21 | 1.5E-64 | 5.31E-47 | 2.99E-54 | 3.03E-55 | 1.19E-52 | 5.17E-05 | 4.06E-38 | 3.38E-39 | 5.96E-22 |
| *LCA* | 1.39E-39 | 6.54E-26 | 9.96E-60 | 6.26E-49 | 5.31E-39 | 2.21E-33 | 1.7E-121 | 7.3E-17 | 2.1E-105 | NA | 1.21E-17 | 2.86E-41 | 1.8E-53 | 7E-103 | 6.8E-33 | 5.31E-73 | 1.8E-166 | 1E-143 | 2.27E-35 | 3.39E-72 | 1.33E-15 | 1.33E-50 | 7.5E-128 | 6.23E-23 |
| *MCA* | 3.7E-122 | 6.09E-98 | 3.1E-36 | 8.86E-28 | 4.89E-21 | 1.11E-14 | 1.68E-63 | 2.15E-98 | 1.35E-60 | 1.21E-17 | NA | 8.6E-65 | 1.89E-90 | 4.17E-23 | 2.47E-22 | 1.29E-16 | 5.05E-10 | 1.25E-10 | 4.42E-65 | 6.08E-13 | 4.04E-08 | 3.29E-12 | 1.02E-06 | 5.12E-33 |
| *MDCA* | 5.06E-53 | 1.06E-23 | 3.43E-49 | 4.96E-22 | 1.4E-19 | 6.17E-12 | 2.64E-70 | 5.07E-43 | 3.4E-172 | 2.86E-41 | 8.6E-65 | NA | 4.5E-130 | 6.23E-47 | 6.78E-27 | 2.14E-33 | 5.52E-23 | 2.13E-23 | 2.8E-119 | 1.11E-22 | 9.19E-07 | 2.51E-17 | 1.33E-15 | 7.67E-52 |
| *UDCA* | 1.8E-112 | 1.52E-65 | 7.07E-68 | 1.73E-35 | 3.16E-28 | 8.86E-19 | 5.88E-95 | 1.8E-113 | 6.87E-96 | 1.8E-53 | 1.89E-90 | 4.5E-130 | NA | 8.38E-62 | 1.91E-32 | 5.14E-31 | 6.06E-35 | 6.29E-29 | 5.4E-103 | 4.77E-26 | 1.96E-09 | 4.3E-17 | 1.09E-19 | 3.94E-50 |
| *GDCA* | 1.83E-42 | 2.2E-29 | 4.4E-141 | 1.3E-118 | 3.72E-91 | 3.68E-86 | 1.6E-155 | 2.01E-27 | 1.63E-88 | 7E-103 | 4.17E-23 | 6.23E-47 | 8.38E-62 | NA | 1.33E-66 | 2.9E-168 | 3.3E-156 | 2.8E-102 | 2.47E-87 | 7E-276 | 5.86E-29 | 2E-114 | 2.1E-158 | 1.27E-65 |
| *GHCA* | 3.89E-31 | 1.83E-43 | 8.8E-156 | 2.1E-176 | 5.6E-146 | 7.2E-127 | 8.66E-15 | 1.02E-49 | 1.54E-21 | 6.8E-33 | 2.47E-22 | 6.78E-27 | 1.91E-32 | 1.33E-66 | NA | 8.43E-29 | 5.61E-37 | 4.67E-52 | 7.7E-109 | 4.94E-80 | 1.3E-184 | 7.47E-59 | 3.89E-65 | 1.3E-110 |
| *GHDCA* | 6.15E-20 | 3.92E-10 | 1.1E-47 | 2.16E-50 | 2.32E-31 | 1.2E-33 | 1.1E-100 | 3.46E-14 | 1.5E-64 | 5.31E-73 | 1.29E-16 | 2.14E-33 | 5.14E-31 | 2.9E-168 | 8.43E-29 | NA | 3.9E-101 | 3.84E-85 | 6.06E-46 | 3E-99 | 1.14E-10 | 8.5E-122 | 1.78E-77 | 3.66E-28 |
| *GLCA* | 8.24E-28 | 1.14E-22 | 8E-110 | 8.11E-78 | 6.12E-66 | 2.54E-52 | 4.82E-67 | 1.08E-17 | 5.31E-47 | 1.8E-166 | 5.05E-10 | 5.52E-23 | 6.06E-35 | 3.3E-156 | 5.61E-37 | 3.9E-101 | NA | 1.2E-127 | 9.57E-50 | 3.95E-96 | 9.2E-17 | 4.33E-52 | 2.1E-218 | 1.45E-32 |
| *GLCA-S* | 5.39E-18 | 2.01E-13 | 2.94E-50 | 1.93E-49 | 2.92E-38 | 1.45E-37 | 3.32E-60 | 5.77E-14 | 2.99E-54 | 1E-143 | 1.25E-10 | 2.13E-23 | 6.29E-29 | 2.8E-102 | 4.67E-52 | 3.84E-85 | 1.2E-127 | NA | 7.51E-28 | 2.95E-69 | 9.44E-20 | 1.85E-62 | 1.5E-120 | 8.13E-18 |
| *GUDCA* | 2.57E-77 | 3.07E-62 | 5.7E-176 | 5.6E-122 | 2.51E-98 | 6.21E-63 | 1.72E-47 | 1.48E-60 | 3.03E-55 | 2.27E-35 | 4.42E-65 | 2.8E-119 | 5.4E-103 | 2.47E-87 | 7.7E-109 | 6.06E-46 | 9.57E-50 | 7.51E-28 | NA | 9.56E-61 | 1.35E-39 | 1.15E-30 | 4.74E-43 | 7.4E-219 |
| *TDCA* | 1.49E-25 | 7.39E-18 | 5.9E-110 | 1.8E-139 | 5.5E-187 | 3E-185 | 1.89E-85 | 1.62E-10 | 1.19E-52 | 3.39E-72 | 6.08E-13 | 1.11E-22 | 4.77E-26 | 7E-276 | 4.94E-80 | 3E-99 | 3.95E-96 | 2.95E-69 | 9.56E-61 | NA | 4.27E-84 | 6.8E-190 | 1.4E-223 | 2.3E-121 |
| *THCA* | 5.6E-11 | 1.49E-19 | 4.11E-71 | 1.2E-109 | 5.1E-179 | 3.1E-168 | 0.00082 | 5.05E-21 | 5.17E-05 | 1.33E-15 | 4.04E-08 | 9.19E-07 | 1.96E-09 | 5.86E-29 | 1.3E-184 | 1.14E-10 | 9.2E-17 | 9.44E-20 | 1.35E-39 | 4.27E-84 | NA | 4.19E-62 | 9.98E-63 | 1.8E-117 |
| *THDCA* | 5.21E-12 | 8.92E-08 | 3.83E-43 | 2.95E-75 | 3.26E-83 | 2.7E-104 | 4.15E-49 | 1.21E-08 | 4.06E-38 | 1.33E-50 | 3.29E-12 | 2.51E-17 | 4.3E-17 | 2E-114 | 7.47E-59 | 8.5E-122 | 4.33E-52 | 1.85E-62 | 1.15E-30 | 6.8E-190 | 4.19E-62 | NA | 1E-123 | 3.27E-67 |
| *TLCA* | 7.83E-21 | 1.69E-14 | 9.3E-110 | 1E-104 | 4.4E-151 | 3.5E-129 | 2.01E-51 | 6.06E-09 | 3.38E-39 | 7.5E-128 | 1.02E-06 | 1.33E-15 | 1.09E-19 | 2.1E-158 | 3.89E-65 | 1.78E-77 | 2.1E-218 | 1.5E-120 | 4.74E-43 | 1.4E-223 | 9.98E-63 | 1E-123 | NA | 2.86E-76 |
| *TUDCA* | 6.65E-41 | 2.0E-33 | 2.1E-131 | 4.6E-149 | 1.4E-223 | 7E-163 | 9.64E-20 | 3.87E-37 | 5.96E-22 | 6.23E-23 | 5.12E-33 | 7.67E-52 | 3.94E-50 | 1.27E-65 | 1.3E-110 | 3.66E-28 | 1.45E-32 | 8.13E-18 | 7.4E-219 | 2.3E-121 | 1.8E-117 | 3.27E-67 | 2.86E-76 | NA |

*Supplementary Table S7.* $log_2$ fold changes, including 95% confidence intervals and p-values for comparisons of all PwMS and matched controls shown in Figure 1A. A p-value ≤0.0125 was considered significant, significant p-values are highlighted with bold text.

| | ***MS vs MC*** | | ***RRMS vs MC*** | | ***PMS vs MC*** | |
|---|---|---|---|---|---|---|
| | ***log₂ FC (95% CI)*** | ***p-value*** | ***log₂ FC (95% CI)*** | ***p-value*** | ***log₂ FC (95% CI)*** | ***p-value*** |
| *CDCA* | -0.332 (-0.662 - -0.003) | 0.048 | -0.205 (-0.435 - 0.025) | 0.081 | -0.396 (-0.857 - 0.064) | 0.092 |
| *CA* | -0.153 (-0.496 - 0.19) | 0.382 | -0.085 (-0.326 - 0.156) | 0.489 | -0.187 (-0.665 - 0.291) | 0.443 |
| *Primary non-conjugated* | -0.322 (-0.638 - -0.006) | 0.046 | -0.219 (-0.440 - 0.001) | 0.051 | -0.373 (-0.814 - 0.069) | 0.098 |
| *GCDCA* | **-0.291 (-0.510 - -0.073)** | **9.06E-03** | **-0.302 (-0.451 - -0.153)** | **7.88E-05** | -0.286 (-0.594 - 0.022) | 0.069 |
| *GCA* | -0.261 (-0.499 - -0.023) | 0.032 | **-0.297 (-0.460 - -0.134)** | **3.76E-04** | -0.243 (-0.578 - 0.093) | 0.156 |
| *TCDCA* | -0.295 (-0.560 - -0.030) | 0.029 | **-0.351 (-0.534 - -0.168)** | **1.83E-04** | -0.267 (-0.639 - 0.105) | 0.159 |
| *TCA* | -0.242 (-0.555 - 0.070) | 0.128 | **-0.332 (-0.549 - -0.114)** | **2.87E-03** | -0.198 (-0.634 - 0.239) | 0.374 |
| *Primary conjugated* | -0.265 (-0.483 - -0.046) | 0.018 | **-0.301 (-0.451 - -0.152)** | **8.40E-05** | -0.246 (-0.555 - 0.062) | 0.118 |
| *DCA* | -0.058 (-0.374 - 0.258) | 0.719 | 0.030 (-0.189 - 0.249) | 0.788 | -0.102 (-0.544 - 0.340) | 0.651 |
| *HCA* | -0.126 (-0.468 - 0.216) | 0.470 | -0.213 (-0.451 - 0.024) | 0.079 | -0.082 (-0.562 - 0.397) | 0.736 |
| *HDCA* | -0.089 (-0.339 - 0.161) | 0.485 | 0.140 (-0.032 - 0.312) | 0.111 | -0.204 (-0.556 - 0.149) | 0.257 |
| *LCA* | -0.165 (-0.410 - 0.081) | 0.188 | -0.003 (-0.172 - 0.165) | 0.968 | -0.245 (-0.590 - 0.099) | 0.163 |
| *MCA* | 0.046 (-0.260 - 0.351) | 0.769 | 0.175 (-0.039 - 0.388) | 0.109 | -0.019 (-0.447 - 0.409) | 0.931 |
| *MDCA* | -0.149 (-0.444 - 0.147) | 0.324 | 0.115 (-0.093 - 0.323) | 0.278 | -0.281 (-0.692 - 0.131) | 0.181 |
| *UDCA* | -0.086 (-0.377 - 0.206) | 0.564 | 0.030 (-0.173 - 0.233) | 0.774 | -0.143 (-0.550 - 0.264) | 0.490 |
| *Secondary non-conjugated* | -0.101 (-0.336 - 0.133) | 0.397 | 0.048 (-0.113 - 0.208) | 0.562 | -0.176 (-0.507 - 0.155) | 0.297 |
| *GDCA* | -0.041 (-0.379 - 0.297) | 0.813 | -0.129 (-0.367 - 0.108) | 0.286 | 0.003 (-0.467 - 0.473) | 0.989 |
| *GHCA* | -0.246 (-0.459 - -0.034) | 0.023 | **-0.259 (-0.403 - -0.114)** | **4.75E-04** | -0.240 (-0.540 - 0.060) | 0.116 |
| *GHDCA* | -0.103 (-0.469 - 0.262) | 0.579 | -0.050 (-0.305 - 0.206) | 0.704 | -0.130 (-0.641 - 0.380) | 0.617 |
| *GLCA* | -0.433 (-0.784 - -0.081) | 0.016 | **-0.359 (-0.606 - -0.113)** | **4.35E-03** | -0.470 (-0.960 - 0.020) | 0.060 |
| *GLCA-S* | 0.030 (-0.208 - 0.268) | 0.803 | 0.030 (-0.133 - 0.193) | 0.717 | 0.030 (-0.305 - 0.366) | 0.859 |
| *GUDCA* | -0.221 (-0.473 - 0.031) | 0.086 | -0.206 (-0.38 - -0.033) | 0.020 | -0.228 (-0.583 - 0.126) | 0.206 |
| *TDCA* | -0.122 (-0.468 - 0.224) | 0.489 | -0.226 (-0.47 - 0.017) | 0.069 | -0.070 (-0.550 - 0.411) | 0.776 |
| *THCA* | -0.161 (-0.526 - 0.204) | 0.386 | -0.252 (-0.508 - 0.003) | 0.053 | -0.116 (-0.626 - 0.395) | 0.657 |
| *THDCA* | -0.148 (-0.522 - 0.227) | 0.439 | -0.305 (-0.571 - -0.039) | 0.025 | -0.069 (-0.59 - 0.452) | 0.795 |
| *TLCA* | -0.193 (-0.501 - 0.116) | 0.221 | **-0.282 (-0.497 - -0.067)** | **0.010** | -0.148 (-0.579 - 0.283) | 0.501 |
| *TUDCA* | -0.192 (-0.505 - 0.122) | 0.230 | -0.265 (-0.483 - -0.047) | 0.017 | -0.155 (-0.594 - 0.284) | 0.489 |
| *Secondary conjugated* | -0.071 (-0.267 - 0.126) | 0.480 | **-0.184 (-0.317 - -0.052)** | **6.59E-03** | -0.014 (-0.291 - 0.264) | 0.923 |
| *Total bile acids* | -0.148 (-0.325 - 0.028) | 0.099 | **-0.220 (-0.339 - -0.101)** | **2.99E-04** | -0.113 (-0.363 - 0.138) | 0.378 |

*Supplementary Table S8.* $\log_2$ fold changes, including 95% confidence intervals and p-values for comparisons of **male** PwMS and matched controls shown in Figure 1B. A p-value ≤0.0125 was considered significant, significant p-values are highlighted with bold text.

| | ***MS vs MC*** | | ***RRMS vs MC*** | | ***PMS vs MC*** | |
|---|---|---|---|---|---|---|
| | ***log₂ FC (95% CI)*** | ***p-value*** | ***log₂ FC (95% CI)*** | ***p-value*** | ***log₂ FC (95% CI)*** | ***p-value*** |
| *CDCA* | -0.469 (-0.989 - 0.051) | 0.077 | -0.316 (-0.712 - 0.08) | 0.118 | -0.546 (-1.26 - 0.171) | 0.136 |
| *CA* | -0.219 (-0.761 - 0.324) | 0.429 | -0.264 (-0.679 - 0.151) | 0.212 | -0.196 (-0.942 - 0.55) | 0.607 |
| *Primary non-conjugated* | -0.470 (-0.814 - -0.125) | 0.065 | -0.432 (-0.689 - -0.175) | 0.040 | -0.488 (-0.97 - -0.007) | 0.150 |
| *GCDCA* | **-0.481 (-0.857 - -0.106)** | **7.66E-03** | **-0.486 (-0.767 - -0.206)** | **1.02E-03** | -0.479 (-1.00 - 0.045) | 0.047 |
| *GCA* | **-0.486 (-0.905 - -0.068)** | **0.012** | **-0.529 (-0.845 - -0.214)** | **6.97E-04** | -0.464 (-1.05 - 0.117) | 0.073 |
| *TCDCA* | -0.435 (-0.93 - 0.061) | 0.023 | **-0.515 (-0.890 - -0.141)** | **1.02E-03** | -0.394 (-1.08 - 0.289) | 0.117 |
| *TCA* | -0.351 (-0.850 - 0.147) | 0.085 | **0.025 (-0.352 - 0.402)** | **7.05E-03** | -0.539 (-1.23 - 0.151) | 0.258 |
| *Primary conjugated* | **-0.424 (-0.964 - 0.116)** | **9.08E-03** | **-0.474 (-0.883 - -0.065)** | **4.25E-04** | -0.399 (-1.15 - 0.35) | 0.063 |
| *DCA* | -0.215 (-0.610 - 0.180) | 0.167 | 0.263 (-0.033 - 0.559) | 0.896 | -0.454 (-1.004 - 0.096) | 0.126 |
| *HCA* | -0.357 (-0.742 - 0.028) | 0.124 | 0.097 (-0.193 - 0.386) | 0.023 | -0.584 (-1.119 - -0.048) | 0.296 |
| *HDCA* | -0.295 (-0.762 - 0.173) | 0.286 | 0.218 (-0.148 - 0.583) | 0.081 | -0.551 (-1.197 - 0.095) | 0.106 |
| *LCA* | -0.364 (-0.831 - 0.104) | 0.069 | 0.205 (-0.153 - 0.564) | 0.512 | -0.649 (-1.29 - -0.007) | 0.033 |
| *MCA* | -0.402 (-0.862 - 0.058) | 0.216 | 0.021 (-0.328 - 0.371) | 0.243 | -0.613 (-1.249 - 0.022) | 0.095 |
| *MDCA* | -0.310 (-0.844 - 0.225) | 0.127 | -0.172 (-0.581 - 0.237) | 0.261 | -0.378 (-1.11 - 0.356) | 0.048 |
| *UDCA* | -0.460 (-0.795 - -0.125) | 0.087 | -0.432 (-0.681 - -0.184) | 0.905 | -0.474 (-0.944 - -0.005) | 0.059 |
| *Secondary non-conjugated* | -0.383 (-0.961 - 0.196) | 0.075 | -0.162 (-0.602 - 0.278) | 0.562 | -0.493 (-1.29 - 0.304) | 0.039 |
| *GDCA* | -0.728 (-1.28 - -0.172) | 0.256 | -0.344 (-0.768 - 0.080) | 0.408 | -0.92 (-1.69 - -0.154) | 0.312 |
| *GHCA* | **-0.028 (-0.404 - 0.347)** | **7.10E-03** | **0.134 (-0.146 - 0.414)** | **6.79E-04** | -0.109 (-0.633 - 0.415) | 0.048 |
| *GHDCA* | -0.447 (-0.845 - -0.049) | 0.194 | -0.394 (-0.693 - -0.095) | 0.471 | -0.474 (-1.03 - 0.079) | 0.225 |
| *GLCA* | **-0.369 (-0.916 - 0.177)** | **0.010** | -0.306 (-0.725 - 0.113) | 0.112 | -0.401 (-1.15 - 0.349) | 0.019 |
| *GLCA-S* | -0.394 (-0.967 - 0.179) | 0.883 | -0.465 (-0.906 - -0.024) | 0.348 | -0.358 (-1.15 - 0.433) | 0.682 |
| *GUDCA* | -0.474 (-1.07 - 0.120) | 0.028 | **-0.479 (-0.936 - -0.022)** | **0.010** | -0.471 (-1.29 - 0.346) | 0.093 |
| *TDCA* | -0.456 (-0.944 - 0.033) | 0.185 | -0.403 (-0.773 - -0.033) | 0.152 | -0.482 (-1.16 - 0.192) | 0.295 |
| *THCA* | -0.439 (-0.935 - 0.057) | 0.178 | -0.579 (-0.954 - -0.203) | 0.039 | -0.369 (-1.06 - 0.319) | 0.375 |
| *THDCA* | -0.342 (-0.62 - -0.064) | 0.118 | -0.334 (-0.539 - -0.129) | 0.040 | -0.346 (-0.738 - 0.046) | 0.258 |
| *TLCA* | -0.470 (-0.969 - 0.030) | 0.068 | -0.398 (-0.777 - -0.018) | 0.033 | -0.506 (-1.20 - 0.184) | 0.161 |
| *TUDCA* | -0.336 (-0.707 - 0.034) | 0.083 | **0.082 (-0.195 - 0.359)** | **2.55E-03** | -0.546 (-1.06 - -0.029) | 0.293 |
| *Secondary conjugated* | -0.460 (-0.805 - -0.115) | 0.057 | -0.464 (-0.721 - -0.207) | 0.021 | -0.458 (-0.941 - 0.025) | 0.154 |
| *Total bile acids* | -0.300 (-0.609 - 0.009) | 0.016 | **-0.269 (-0.497 - -0.040)** | **1.41E-03** | -0.316 (-0.750 - 0.119) | 0.084 |

*Supplementary Table S9*. $\log_2$ fold changes, including 95% confidence intervals and p-values for comparisons of **female** PwMS and matched controls shown in Figure 1B. A p-value ≤0.0125 was considered significant, significant p-values are highlighted with bold text.

| | ***MS vs MC*** | | ***RRMS vs MC*** | | ***PMS vs MC*** | |
|---|---|---|---|---|---|---|
| | ***FC (95% CI)*** | ***p-value*** | ***FC (95% CI)*** | ***p-value*** | ***FC (95% CI)*** | ***p-value*** |
| *CDCA* | -0.195 (-0.582 - 0.191) | 0.321 | -0.094 (-0.316 - 0.129) | 0.410 | -0.246 (-0.798 - 0.305) | 0.381 |
| *CA* | -0.087 (-0.487 - 0.313) | 0.669 | 0.094 (-0.140 - 0.328) | 0.431 | -0.178 (-0.747 - 0.392) | 0.540 |
| *Primary non-conjugated* | -0.174 (-0.542 - 0.195) | 0.356 | -0.041 (-0.255 - 0.172) | 0.704 | -0.240 (-0.766 - 0.287) | 0.372 |
| *GCDCA* | -0.113 (-0.369 - 0.143) | 0.387 | -0.172 (-0.317 - -0.028) | 0.019 | -0.083 (-0.452 - 0.285) | 0.658 |
| *GCA* | -0.040 (-0.319 - 0.239) | 0.779 | -0.107 (-0.264 - 0.051) | 0.183 | -0.006 (-0.408 - 0.395) | 0.975 |
| *TCDCA* | -0.104 (-0.414 - 0.206) | 0.510 | -0.172 (-0.349 - 0.005) | 0.057 | -0.070 (-0.515 - 0.374) | 0.756 |
| *TCA* | -0.050 (-0.413 - 0.313) | 0.786 | -0.148 (-0.359 - 0.063) | 0.170 | -0.002 (-0.520 - 0.516) | 0.995 |
| *Primary conjugated* | -0.069 (-0.326 - 0.188) | 0.598 | -0.138 (-0.283 - 0.006) | 0.060 | -0.034 (-0.404 - 0.335) | 0.855 |
| *DCA* | 0.235 (-0.134 - 0.605) | 0.212 | 0.035 (-0.177 - 0.247) | 0.747 | 0.335 (-0.193 - 0.864) | 0.213 |
| *HCA* | 0.172 (-0.228 - 0.572) | 0.398 | 0.048 (-0.182 - 0.277) | 0.686 | 0.235 (-0.338 - 0.807) | 0.421 |
| *HDCA* | 0.037 (-0.257 - 0.330) | 0.807 | 0.016 (-0.150 - 0.182) | 0.850 | 0.047 (-0.374 - 0.468) | 0.828 |
| *LCA* | 0.028 (-0.261 - 0.316) | 0.852 | -0.104 (-0.267 - 0.060) | 0.215 | 0.093 (-0.322 - 0.508) | 0.660 |
| *MCA* | 0.386 (0.011 - 0.761) | 0.044 | 0.132 (-0.078 - 0.342) | 0.218 | 0.513 (-0.024 - 1.05) | 0.061 |
| *MDCA* | 0.067 (-0.277 - 0.411) | 0.704 | 0.025 (-0.177 - 0.227) | 0.809 | 0.088 (-0.402 - 0.577) | 0.726 |
| *UDCA* | 0.230 (-0.109 - 0.570) | 0.184 | 0.038 (-0.159 - 0.235) | 0.703 | 0.327 (-0.159 - 0.812) | 0.187 |
| *Secondary non-conjugated* | 0.134 (-0.141 - 0.409) | 0.341 | 0.013 (-0.142 - 0.169) | 0.866 | 0.194 (-0.202 - 0.589) | 0.337 |
| *GDCA* | 0.228 (-0.166 - 0.621) | 0.256 | -0.086 (-0.317 - 0.144) | 0.462 | 0.385 (-0.175 - 0.945) | 0.178 |
| *GHCA* | -0.032 (-0.282 - 0.217) | 0.799 | -0.085 (-0.224 - 0.055) | 0.234 | -0.006 (-0.365 - 0.353) | 0.972 |
| *GHDCA* | 0.176 (-0.25 - 0.602) | 0.417 | 0.063 (-0.185 - 0.311) | 0.620 | 0.233 (-0.375 - 0.840) | 0.453 |
| *GLCA* | -0.138 (-0.548 - 0.272) | 0.509 | **-0.374 (-0.613 - -0.135)** | **0.002** | -0.020 (-0.604 - 0.564) | 0.947 |
| *GLCA-S* | 0.089 (-0.191 - 0.368) | 0.533 | -0.074 (-0.231 - 0.083) | 0.356 | 0.170 (-0.232 - 0.572) | 0.406 |
| *GUDCA* | 0.005 (-0.290 - 0.300) | 0.973 | -0.019 (-0.187 - 0.149) | 0.827 | 0.017 (-0.406 - 0.440) | 0.937 |
| *TDCA* | 0.125 (-0.277 - 0.528) | 0.541 | -0.146 (-0.382 - 0.090) | 0.225 | 0.261 (-0.311 - 0.833) | 0.371 |
| *THCA* | 0.071 (-0.360 - 0.502) | 0.745 | -0.040 (-0.282 - 0.202) | 0.746 | 0.127 (-0.491 - 0.745) | 0.687 |
| *THDCA* | 0.179 (-0.255 - 0.612) | 0.419 | -0.130 (-0.389 - 0.128) | 0.322 | 0.333 (-0.283 - 0.949) | 0.289 |
| *TLCA* | 0.071 (-0.287 - 0.429) | 0.699 | -0.161 (-0.369 - 0.047) | 0.130 | 0.186 (-0.324 - 0.697) | 0.474 |
| *TUDCA* | 0.056 (-0.309 - 0.420) | 0.764 | 0.048 (-0.162 - 0.258) | 0.651 | 0.059 (-0.463 - 0.581) | 0.823 |
| *Secondary conjugated* | 0.159 (-0.072 - 0.389) | 0.176 | -0.100 (-0.228 - 0.028) | 0.125 | 0.288 (-0.044 - 0.621) | 0.089 |
| *Total bile acids* | 0.045 (-0.163 - 0.253) | 0.670 | -0.106 (-0.220 - 0.008) | 0.069 | 0.121 (-0.180 - 0.421) | 0.430 |

*Supplementary Table S10.* Hazard ratios, including 95% confidence intervals and p-values for the Cox regression of bile acids association with reaching EDSS milestone 4, stratified based sex. A p-value ≤0.0125 was considered significant, significant p-values are highlighted with bold text.

| | ***Males*** | | ***Females*** | |
|---|---|---|---|---|
| | **HR (95% CI)** | **p-value** | **HR (95 % CI)** | **p-value** |
| *CDCA* | 1.01 (0.839 - 1.22) | 0.899 | 1.01 (0.907 - 1.12) | 0.904 |
| *CA* | 0.979 (0.821 - 1.17) | 0.811 | 1.02 (0.927 - 1.12) | 0.708 |
| *Primary non-conjugated* | 0.993 (0.822 - 1.20) | 0.940 | 1.02 (0.920 - 1.13) | 0.699 |
| *GCDCA* | 1.19 (0.957 - 1.49) | 0.117 | **1.26 (1.06 - 1.50)** | **0.008** |
| *GCA* | 1.13 (0.948 - 1.34) | 0.176 | 1.19 (1.02 - 1.39) | 0.029 |
| *TCDCA* | 1.17 (0.98 - 1.40) | 0.083 | 1.16 (1.01 - 1.34) | 0.032 |
| *TCA* | 1.15 (0.987 - 1.35) | 0.072 | 1.10 (0.976 - 1.25) | 0.114 |
| *Primary conjugated* | 1.15 (0.957 - 1.39) | 0.133 | **1.24 (1.05 - 1.48)** | **0.012** |
| *DCA* | 1.05 (0.885 - 1.25) | 0.574 | 1.07 (0.950 - 1.20) | 0.279 |
| *HCA* | 0.929 (0.774 - 1.11) | 0.427 | 0.999 (0.909 - 1.10) | 0.988 |
| *HDCA* | 1.05 (0.848 - 1.30) | 0.651 | 1.13 (0.987 - 1.28) | 0.078 |
| *LCA* | 1.18 (0.908 - 1.52) | 0.219 | 1.15 (1.01 - 1.31) | 0.031 |
| *MCA* | 1.03 (0.853 - 1.25) | 0.747 | 1.01 (0.897 - 1.13) | 0.915 |
| *MDCA* | 1.18 (0.941 - 1.47) | 0.153 | 1.06 (0.942 - 1.19) | 0.332 |
| *UDCA* | 1.14 (0.948 - 1.37) | 0.164 | 1.03 (0.903 - 1.17) | 0.688 |
| *Secondary non-conjugated* | 1.07 (0.864 - 1.34) | 0.518 | 1.08 (0.944 - 1.23) | 0.271 |
| *GDCA* | 1.11 (0.925 - 1.33) | 0.263 | 1.11 (0.986 - 1.25) | 0.085 |
| *GHCA* | 1.17 (0.926 - 1.49) | 0.184 | 1.17 (1.01 - 1.35) | 0.032 |
| *GHDCA* | **1.23 (1.05 - 1.44)** | **0.010** | 1.03 (0.942 - 1.13) | 0.506 |
| *GLCA* | 1.09 (0.933 - 1.28) | 0.272 | 1.10 (1.00 - 1.22) | 0.051 |
| *GLCAS* | 1.19 (0.937 - 1.50) | 0.156 | 1.13 (0.999 - 1.28) | 0.052 |
| *GUDCA* | 1.20 (0.963 - 1.49) | 0.105 | 1.11 (0.967 - 1.28) | 0.138 |
| *TDCA* | 1.12 (0.948 - 1.33) | 0.180 | 1.09 (0.973 - 1.23) | 0.133 |
| *THCA* | 1.23 (0.975 - 1.55) | 0.081 | 1.05 (0.949 - 1.16) | 0.338 |
| *THDCA* | 1.13 (0.973 - 1.31) | 0.111 | 1.07 (0.961 - 1.19) | 0.215 |
| *TLCA* | 1.12 (0.943 - 1.34) | 0.190 | 1.15 (1.02 - 1.30) | 0.023 |
| *TUDCA* | 1.25 (1.03 - 1.52) | 0.025 | 1.02 (0.910 - 1.15) | 0.701 |
| *Secondary conjugated* | 1.19 (0.951 - 1.49) | 0.128 | **1.23 (1.05 - 1.44)** | **0.009** |
| *Total bile acids* | 1.17 (0.944 - 1.45) | 0.151 | 1.25 (1.04 - 1.51) | 0.015 |

*Supplementary Table S11*. Hazard ratios, including 95% confidence intervals and p-values for the Cox regression of bile acids association with reaching EDSS milestone 6.

| | ***HR (95% CI)*** | ***p-value*** |
|---|---|---|
| *CDCA* | 1.03 (0.913 - 1.15) | 0.662 |
| *CA* | 1.00 (0.887 - 1.13) | 0.969 |
| *Primary unconjugated* | 0.999 (0.869 - 1.15) | 0.986 |
| *GCDCA* | 1.08 (0.891 - 1.30) | 0.443 |
| *GCA* | 1.15 (0.981 - 1.34) | 0.085 |
| *TCDCA* | 1.07 (0.930 - 1.23) | 0.342 |
| *TCA* | 1.11 (0.984 - 1.26) | 0.087 |
| *Primary conjugated* | 1.12 (0.939 - 1.33) | 0.213 |
| *DCA* | 1.08 (0.943 - 1.24) | 0.259 |
| *HCA* | 0.980 (0.872 - 1.10) | 0.740 |
| *HDCA* | 1.05 (0.898 - 1.23) | 0.545 |
| *LCA* | 1.10 (0.932 - 1.29) | 0.263 |
| *MCA* | 1.05 (0.920 - 1.19) | 0.477 |
| *MDCA* | 1.03 (0.893 - 1.19) | 0.665 |
| *UDCA* | 1.01 (0.863 - 1.18) | 0.901 |
| *Secondary unconjugated* | 1.07 (0.908 - 1.26) | 0.415 |
| *GDCA* | 1.09 (0.955 - 1.25) | 0.198 |
| *GHCA* | 1.14 (0.953 - 1.37) | 0.149 |
| *GHDCA* | 1.04 (0.946 - 1.15) | 0.388 |
| *GLCA* | 1.01 (0.913 - 1.12) | 0.812 |
| *GLCAS* | 1.07 (0.918 - 1.24) | 0.404 |
| *GUDCA* | 1.09 (0.910 - 1.30) | 0.350 |
| *TDCA* | 1.11 (0.973 - 1.25) | 0.123 |
| *THCA* | 1.13 (0.990 - 1.29) | 0.067 |
| *THDCA* | 1.06 (0.938 - 1.20) | 0.339 |
| *TLCA* | 1.07 (0.937 - 1.23) | 0.302 |
| *TUDCA* | 1.05 (0.916 - 1.20) | 0.489 |
| *Secondary conjugated* | 1.13 (0.944 - 1.35) | 0.183 |
| *Total bile acids* | 1.15 (0.942 - 1.41) | 0.168 |
